# Extending Genome-Wide Association Studies to admixed cohorts with high degrees of relatedness

**DOI:** 10.1101/2025.05.27.25328444

**Authors:** Taotao Tan, Alejandra Vergara-Lope, José Jaime Martínez-Magaña, Nirav N. Shah, Kai Yuan, Jaime Berumen, Jesus Alegre-Díaz, Pablo Kuri-Morales, Roberto Tapia-Conyer, Joel Gelenter, Janitza L. Montalvo-Ortiz, Wei Zhou, Jason M. Torres, Elizabeth G. Atkinson

## Abstract

Recently admixed populations comprise a large portion of the human population worldwide, but are often excluded from Genome-Wide Association Studies (GWAS) due to analytic challenges. Our group has previously developed a local ancestry informed generalized linear model based method, *Tractor*, for GWAS in admixed samples, which produces accurate ancestry-specific effect sizes and boosts discovery power to identify ancestry-enriched loci. *Tractor* has been instrumental for elucidating the genetic architecture of complex traits across admixed cohorts, however it operates under an assumption of unrelated samples. As biobanks and other large-scale data sources continue to grow, increasing numbers of closely or cryptically related admixed samples are included. This brings new statistical challenges in conducting GWAS and motivates the timely development of novel tools that can model admixture in various cohort settings. Here, we propose a novel mixed model method, *Tractor-Mix*, that allows for well-calibrated association studies in datasets containing admixed samples with high degrees of relatedness. Similar to *Tractor*, our method conducts genetic association tests by leveraging local ancestry to produce more accurate effect sizes and boost power under heterogeneity while effectively controlling false positives. Extensive simulations show this enhanced method is competitive with other state-of-the-art approaches that do not produce ancestry-specific results. Empirical testing of *Tractor-Mix* on multiple cohorts, including admixed samples from the UK Biobank and Mexico City Prospective Study, highlight the value of the method, identifying ancestry-specific associations. In summary, *Tractor-Mix* is a powerful association framework that extends the capabilities of current models and will facilitate the inclusion of admixed samples in large-scale GWAS.

## Introduction

Genome-wide association studies (GWAS) have revolutionized our understanding of the genetic basis of complex traits and identified thousands of loci associated with complex diseases^1–3^. However, the majority of GWAS have been conducted on homogeneous European populations, which can lead to non-generalizable findings^4–7^. Despite making up more than a third of the US populace and being the fastest growing demographic^8^, admixed populations only comprise less than 7% of all published GWAS^9^. Gaps in sample recruitment contribute to this issue, but even when data exists, admixed individuals have historically been excluded due to the scarcity of methods and pipelines to effectively account for admixture. A major concern in analyzing global populations is the confounding effect of population structure. As a workaround, samples are typically grouped by ancestry, and GWAS is conducted independently for each subset of the cohort. While this approach helps mitigate confounding effects, it often leads to the exclusion of samples with admixed backgrounds, which might lead to reduced power^10–15^.

Numerous ongoing efforts are actively working to build more representative biobanks or aggregated collections that include higher numbers of individuals with genetic admixture. Complementing this is targeted cohort recruitment occurring on smaller scales internationally. The growing diversity in genomic data represents a significant advancement for the field, enhancing the breadth of health research. However, this increased complexity requires careful consideration in analysis. As large datasets incorporate ever more samples, instances of genetic relatedness become increasingly common. Targeted collections in non-Western settings may also have higher levels of admixture or endogamy than the Western populations around which traditional GWAS tools were designed^16–24^, or be more likely to employ household-based recruitment strategies^16^. For example, in the Mexico City Prospective Study, 70% of participants have at least a third-degree relative within the study^25^. To maximize the utility of expanding genomic resources and ensure that discoveries are relevant to individuals of all ancestries, it is crucial to develop appropriate analytical strategies that can both accurately include admixed individuals and properly account for relatedness.

We previously developed a GWAS framework for admixed populations, called *Tractor*, which allows for well-calibrated study of admixed cohorts and produces ancestry-specific summary statistics^26^. Admixed populations inherit genomic segments from multiple source populations, which can be inferred with local ancestry deconvolution. By allocating risk alleles to each local ancestry background, *Tractor* conducts regression analysis on ancestry-specific genotype dosages, conditioning (optionally) on local ancestry status and other covariates. The summary statistics obtained from *Tractor* consist of accurate ancestry-level effect size estimates and *P* values, allowing for more refined ancestral consideration in downstream analyses such as polygenic-risk score calculations, meta-analysis, and fine mapping efforts^27^. *Tractor* can also boost power to discover loci that show differential effect sizes, whether causal or marginal, among source populations, and reduces the credible set of variants within association hits. However, *Tractor* - and the broader class of generalized linear models (GLMs) to which it belongs - assumes only unrelated samples are included in analyses. As standard GLM regression-based GWAS requires samples to be independent, large numbers of individuals harboring relatedness would typically be excluded. This results in loss of statistical power due to a reduced sample size. A more principled approach to handle sample relatedness is to utilize a generalized linear mixed model (GLMM) framework, where pairwise relationships are captured in a genetic relationship matrix (GRM). Programs such as *GMMAT*^28^, *SAIGE*^29^, and *BOLT-LMM*^30^ effectively implement the mixed effect model approach, allowing for scalable GWAS analysis. However, most of the developed approaches are designed for homogeneous populations and do not explicitly account for genetic admixture.

In the method proposed here, we inherit the intuition from the original *Tractor* model and further extend its capacity to admixed samples with relatedness. This new approach, called *Tractor-Mix*, takes the estimated GRM as input and runs a mixed model for each variant (similar to *GMMAT*), while accounting for each allele’s local ancestry background (similar to *Tractor*). Unlike traditional GWAS, *Tractor-Mix* utilizes ancestry-specific genotypes as independent variables and estimates effect sizes and *P* values for each ancestry. Here, we describe our new mixed model framework (Extended Data Figure 1) and extensively evaluate its performance with a simulated dataset, benchmarking against other leading tools. We demonstrate that *Tractor-Mix* has a well-controlled false positive rate. Similar to *Tractor*, it is tuned to detect ancestry-enriched signals, such that it gains statistical power under larger effect size heterogeneity while losing some power if effects are similar across ancestries. *Tractor-Mix* is additionally able to produce reliable effect size estimates at the ancestry-level, which may prove vital for downstream use. To ensure our new model performed well on empirical data, we applied *Tractor-Mix* to admixed individuals of African and European descent from the UK Biobank and Yale-Penn cohort, and of Indigenous American and European descent from the Mexico City Prospective Study. *Tractor-Mix* was able to replicate known hits for multiple traits, as well as detected several loci uniquely identified on specific ancestry backgrounds.

## Results

### *Tractor-Mix* has a well-controlled false positive rate

For a 2-way admixed cohort (in this example, African-European admixture, or AFR-EUR), *Tractor-Mix* performs a 2 degree of freedom (d.o.f.) score test to jointly test the null hypothesis that the effect sizes for both ancestries are zero: β_*A*_ = β_*E*_ = 0 (here, and throughout the article, β_*A*_, β_*E*_ represent effect sizes for two ancestral populations, which can be generalized beyond AFR and EUR). We evaluated false positives for both continuous and dichotomous phenotypes for a simulated admixed dataset with a high degree of relatedness. The phenotype is generated according to a known admixture proportion and kinship matrix (Supplementary Figure 1). However, such information is often unavailable in empirical biobank settings and requires approximations from the genotype matrix. To characterize how admixture proportion estimation and kinship estimation impact false positives, we designed the following eight models (See *Methods*):

1. Standard linear/logistic regression
2. Original (linear/logistic regression) *Tractor*
3. *GMMAT* with a standard estimator for principal components (PCs) and GRM
4. *Tractor-Mix* with a standard estimator for PCs and GRM
5. *GMMAT* with PCs and GRM estimated via PC-AiR and PC-Relate
6. *Tractor-Mix* with PCs and GRM estimated via PC-AiR and PC-Relate
7. *GMMAT* with true admixture proportions and kinship matrix
8. *Tractor-Mix* with true admixture proportions and kinship matrix

In this simulation setup, Model 1, 3, 5, 7 use the total risk allele counts, while the Tractor models - Models 2, 4, 6, 8 - use ancestry-specific allele counts. Models 1 and 2 used standard linear/logistic regression, while Models 3-8 used a linear/logistic mixed model. Models 3 and 4 utilized the first PC, computed via eigen-decomposition of the linkage disequilibrium (LD) matrix 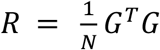, along with the standard GRM, defined as 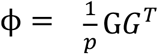(*N* stands for sample size, *p* stands for number of variants, genotype matrix *G* is standardized). Models 5 and 6, on the other hand, employed PC-AiR and PC-Relate^31,32^, which disentangle sample relatedness to population level (captured by PC), and family level (captured by GRM). Through sample partitioning, PC-AiR and PC-Relate tend to recover the true population structure and kinship, and potentially help GWAS. Models 7 and 8 include the simulated – i.e. perfectly estimated – GRMs.

We observe that using a standard linear/logistic regression (Models 1 and 2) without controlling for sample relatedness can yield large counts of false positives, as expected. This is true for both standard GWAS and *Tractor* GWAS (Figure 1). For continuous traits, using a mixed model can alleviate this issue, although small amounts of type I error inflation are still observed when the admixture proportions and kinship matrix are estimated. To assess the impact of various strategies for computing the GRM, we compared the standard estimator (Models 3 and 4: standard PC, standard GRM) with an estimator designed for admixed populations (Models 5 and 6: PC-AiR, PC-Relate). Standard estimators often fail to distinguish relatedness arising from population versus family structure, whereas PC-AiR and PC-Relate can better disentangle these effects. Consequently, GRMs derived from PC-Relate are often sparser than those derived from standard estimators, enabling more efficient algebraic operations during GWAS and, therefore, faster computations^33^. Surprisingly, although PC-AiR and PC-Relate more accurately recover the true admixture proportions and kinship matrix, we found little difference in false positive rates across different GRM generation strategies, consistent with the simulation studies conducted by Gogarten *et al*.^33^. Interestingly, using estimated kinship and admixture proportions can induce small amounts of false positives for continuous phenotypes, but not for dichotomous phenotypes (Figure 1 & Supplementary Figure 2). Finally, if we pass the true admixture proportions and kinship matrix to *Tractor-Mix* (Models 7 & 8), we have very well controlled type I error, indicating well calibrated behavior for the proposed 2 d.o.f. test.

**Figure 1.**
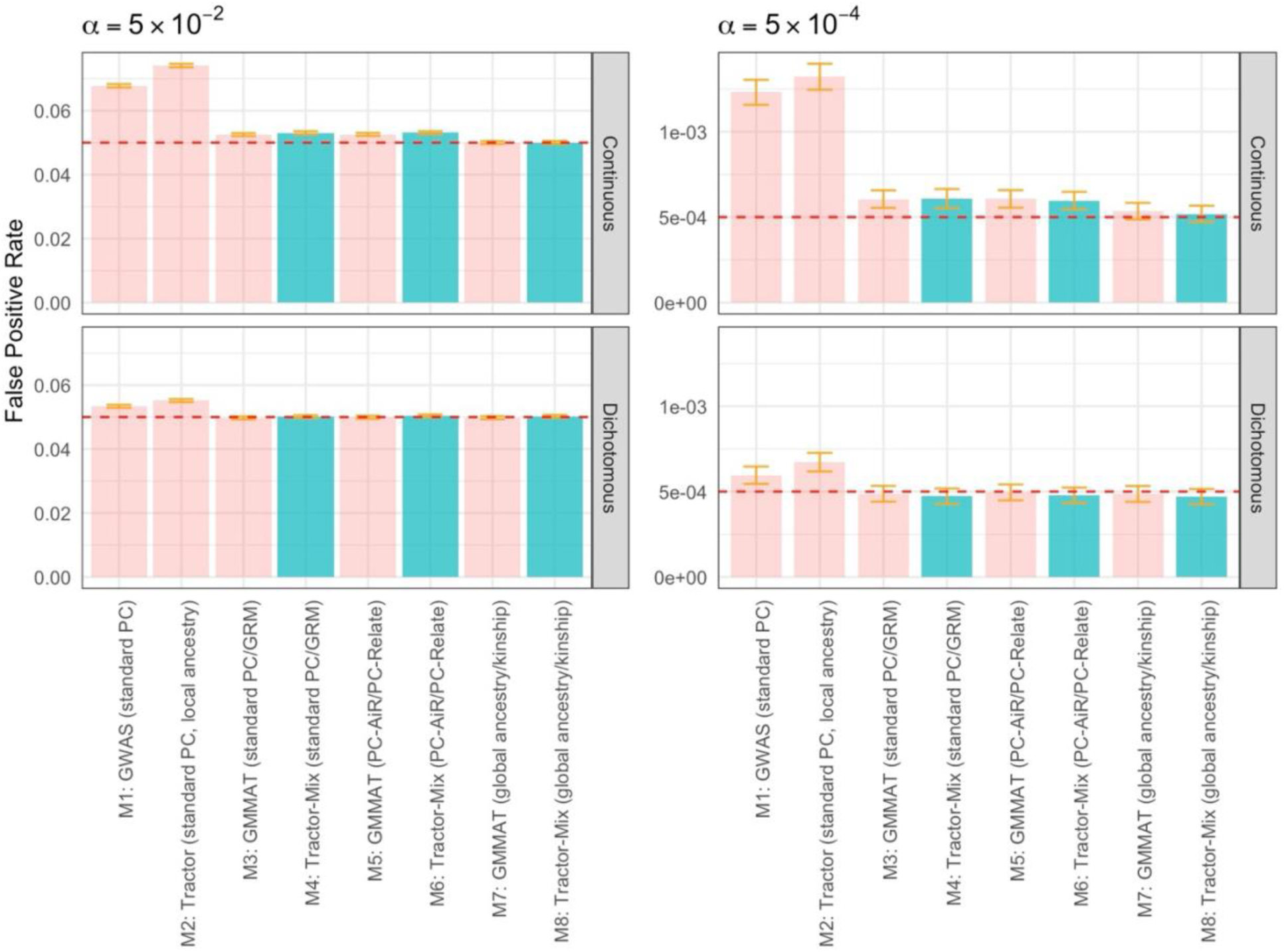
*Tractor-Mix* has a well-controlled false positive rate for admixed cohorts with sample relatedness. False positive rates were computed across our 8 models resulting in alpha significance cutoffs of (**a**) *α* = 5 × 10^−2^ and (**b**) *α* = 5 × 10^−4^. Error bars represent the 95% confidence interval with standard error calculated as 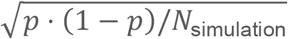. Using linear/logistic regression without considering sample relatedness can results in inflated type I error (pink). Using estimated admixture proportions and kinship matrices (green, blue) can result in a small amount of inflation for continuous traits, but not dichotomous traits. Using the true admixture proportion and kinship (purple, data generation process) results in well-controlled false positive rates. Applying traditional mixed effect based GWAS on admixed samples does not necessarily lead to false positive rate inflations. Note that the joint *P* values are used for false positive evaluation. Darker shaded columns (M4, M6, M8) are derived from Tractor-Mix.

We further tested these 8 models on simulated data with different parameter settings— varying the minor allele frequencies (MAFs), effect size for global admixture proportion, and variance components—and observe similar patterns (See *Methods*, Supplementary Figure 2). Larger scale simulations were implemented for both continuous and dichotomous traits, and we found no false positive inflation for common variants (MAF > 0.05) (Supplementary Table 1). We note that using standard 1 d.o.f. mixed model based GWAS (Models 3, 5, 7) can still yield well-controlled false positives for admixed cohorts in this simulation. Prior work, however, has demonstrated that GRMs alone (as implemented with SAIGE) are insufficient to control for stratification in cohorts with genetic data spanning multiple areas of the globe^34^.

Similar to *GMMAT, Tractor-Mix* might suffer when there are low allele counts and/or a very imbalanced case/control ratio^29^, resulting in increased false positives rates. The procedure of allocating risk allele dosages can exacerbate this issue, resulting in lower ancestry-specific allele counts relative to the total. We explore this issue empirically in Supplementary Figure 3. To address this concern, we implement a strategy in *Tractor-Mix* of dropping terms for particular ancestry-specific genotype dosages when there are too low allele counts (default AC > 50) before computing the *P* value (See *Methods*). This fixes the issue while still allowing for computation of ancestry-specific summary statistics for the ancestries in which the variant is enriched. Variants for which this procedure occurred are tracked in the output so that users are aware which model was utilized and could easily filter to variants for which the full model was run, if that is preferred.

### Evaluating the statistical power and effect size estimates of *Tractor-Mix*

We quantified the statistical power of *Tractor-Mix* for admixed populations with extensive simulations under the Pritchard-Stephens-Donnelly model^35,36^. As admixed populations inherit genetic material from multiple source populations, it is likely that alleles residing in different local ancestries will have different patterns of linkage disequilibrium (LD) and MAF^37–39^. Due to these differences, marginal effect size heterogeneity can be induced, even when causal variants are shared across ancestries. GWAS predominantly leverages the marginal effects of tagging SNPs to identify risk regions, as the causal risk alleles may not be directly genotyped or well captured by the imputation panel due to their low frequencies. In our simulations, the effect sizes can be interpreted interchangeably as marginal or causal effects. Here, we benchmark the landscape of power of our novel model under a variety of effect sizes among admixed populations.

To examine the impact of cross-ancestry effect size heterogeneity on the power of *Tractor-Mix*, we generated a two-way admixed cohort with an average of 80% African (AFR) global genetic ancestry and 20% European (EUR) global genetic ancestry, and fixed the ratio of ancestry-specific effect sizes (β_*A*_: β_*E*_ representing the effect size ratio for AFR and EUR effects, respectively) in each of our simulations. When setting variant effects present in one ancestry while absent in the other, we found substantial power gains with Tractor-Mix (Figure 2a, 2g). This pattern is especially pronounced when variant effects are present in EUR but not AFR. The intuition of this phenomenon is that ancestry-specific signals in the rarer ancestry get diluted by haplotypes from the majority ancestry in standard GWAS, and with local ancestry inference, we recover the proportional sample size for such signals. When we set β_*A*_ = 2β_*E*_ (Figure 2c), we observed similar statistical power for *Tractor-Mix* and *GMMAT*, indicating moderate effect size heterogeneity may not benefit *Tractor-Mix* as much. When we set β_*A*_ = β_*E*_ (Figure 2e), *Tractor-Mix* is less powerful than the standard mixed effect model GWAS. This is expected, as when the component ancestries are identical, resolving the local ancestry of alleles does not add resolution, and will soak up power due to the sparser partitioned dosages. Previous genetic correlation studies have observed conflicting results regarding the level effect size heterogeneity across ancestries in empirical data^40,41^. Whether *Tractor-Mix* will be expected to broadly improve statistical power would depend on the genetic architecture of the trait and the composition of the cohort. We reiterate that GWAS signals are overwhelmingly driven by the marginal effects at tagging loci rather than the causal loci themselves, such that some degree of heterogeneity is expected to be present at many loci across the genome^1,37,42–45^, not only at loci with true causal effect differences across ancestries.

**Figure 2.**
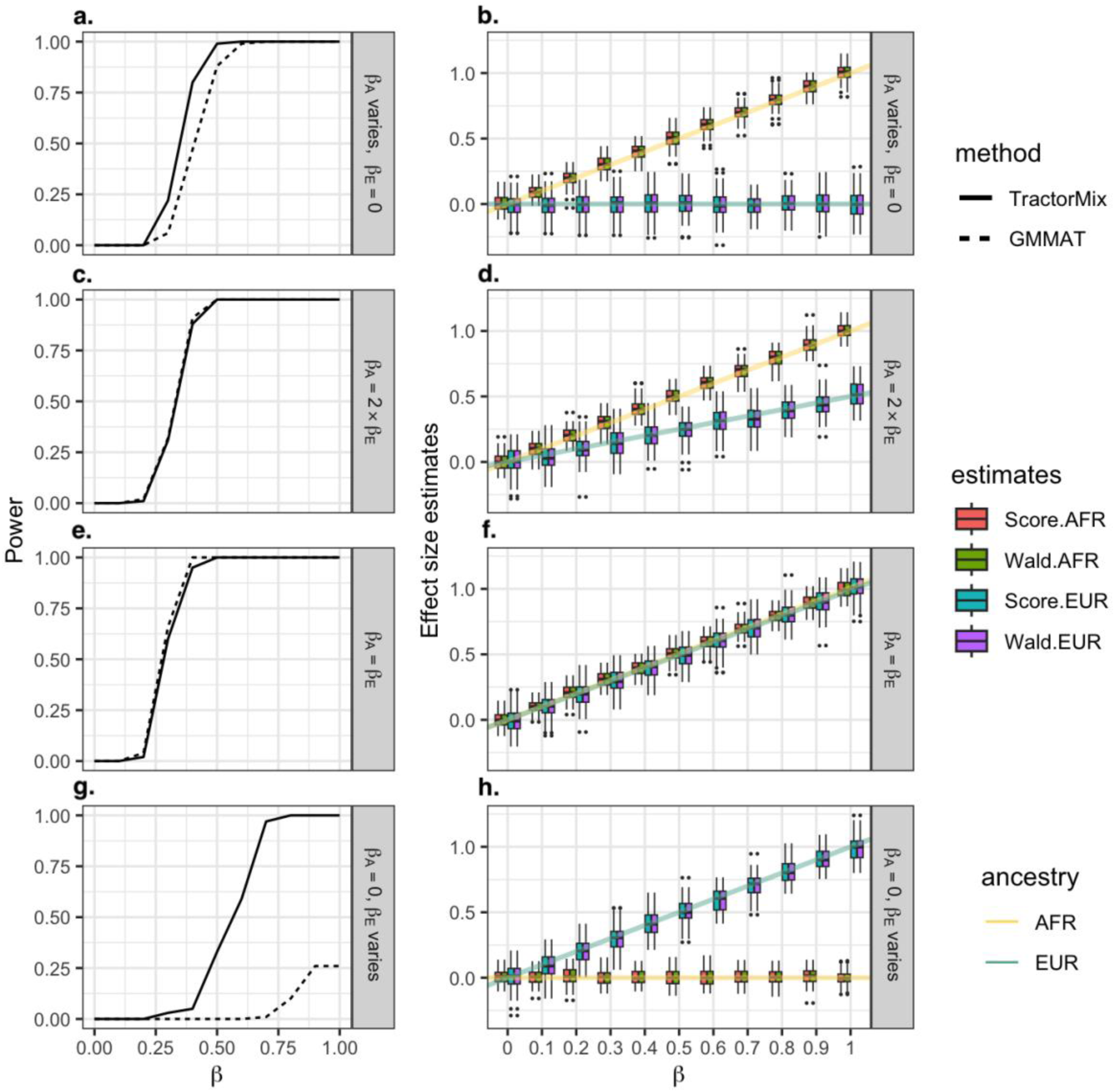
Evaluation of effect size estimation and power for *Tractor-Mix*. The left column shows the statistical power of *Tractor-Mix* and *GMMAT* in different scenarios. *Tractor-Mix* is more powerful with large effect size heterogeneity, equivalent with modest heterogeneity, and is less powerful if effects are identical. Note that the simulation uses the joint *P* value (2 d.o.f.) for power comparisons. The right column compares the accuracy of effect size estimates produced from two strategies (Wald and score tests). The Wald test directly fits the full mixed effect model, while the score test approximates the effect sizes from the null model. The lines are the true effect sizes for each ancestry, with yellow representing AFR and blue representing EUR. As both tests yield accurate and unbiased effect estimates and the score test is more scalable, we implement the score test in the primary *Tractor-Mix* implementation to quickly compute ancestry specific effect sizes across the genome.

While evaluating statistical power, we also checked the effect size estimates for each simulation. We compared two approaches: a Wald test and a score test approximation approach on both AFR and EUR tracts. The score test approximation approach is more suitable for genome-wide scanning, as the null model is pre-computed and is shared across all variants, making it much more computationally efficient and scalable. Although the procedure of the score test never fits a full model, the effect size can still be approximated (see *Methods*). On the other hand, the Wald test directly fits the mixed model and tends to produce more robust effect size estimates. However, it is extremely computationally demanding, making it suitable for testing a more limited number of variants. Across a range of genetic models, we find that the effect size estimates from the Wald test and the score test are fairly similar (Figure 2). Given computational considerations, we therefore generally recommend estimation of effect sizes with the score test. We note that this strategy can have some small downward bias for dichotomous phenotypes (Extended Data Figure 2), similar to what has been reported in GMMAT^28^. Nevertheless, the score test provides a handy approach to estimate effect sizes for each ancestry at the genome-wide scale. In sum, across a wide range of effect size heterogeneities, we found that *Tractor-Mix* can produce unbiased estimates for group-level, ancestry-specific effect sizes for continuous phenotypes.

Another core feature of *Tractor* is its capability of producing ancestry-specific *P* values, which can be used to identify ancestry-enriched GWAS hits, or hits unique to a given background. Under the mixed effect model framework, we can approximate ancestry specific *P* values via the score test, as described above and in the *Methods*. We further examined the concordance of *P* values generated from the Wald test and the score test (Extended Data Figure 3). Under the null hypothesis, we found that the score test approximation method produced very similar *P* values to the Wald test, indicating no type 1 error inflation is introduced. Under the alternative hypothesis, we found that score test approximation typically is more significant for dichotomous phenotypes while less significant for continuous phenotypes. Despite this slight difference in the *P* values produced, the score approach provides an efficient way to obtain genome-wide *P* values without fitting the full model, which significantly improves computational feasibility for large-scale analyses. Should users prefer to obtain *P* values for top loci from the Wald test, we recommend a two-stage approach, with an initial genome-wide score scan (as implemented by default in the *Tractor-Mix* software), and a subsequent Wald test of loci passing a significance threshold. This is a scalable strategy to obtain reliable genome-wide *P* values, with refined Wald *P* values key loci while saving computational expenditures on irrelevant variants. Code for both procedures is freely available on the *Tractor-Mix* Github.

### Empirical examinations of *Tractor-Mix*

#### UK Biobank

To ensure good performance on real data, we applied *Tractor-Mix* to 9,220 2-way admixed AFR-EUR samples from the UK Biobank^46^. The first PC had a high correlation (0.998) with the inferred global ancestry (Supplement Figure 4), indicating consistent estimates of global ancestry states. KING-robust^47^ analysis identified 315 pairs of 1^st^ degree relatedness, and 489 pair of 2^nd^ degree relatedness in these samples. Using PC-AiR and PC-Relate as fixed and random effects, we applied *Tractor-Mix* and *GMMAT* to 3 phenotypes including total cholesterol (Figure 3a-3d), sickle cell anemia (Figure 3e-3h), and low-density lipoprotein (LDL) cholesterol (Supplement Figure 5).

**Figure 3.**
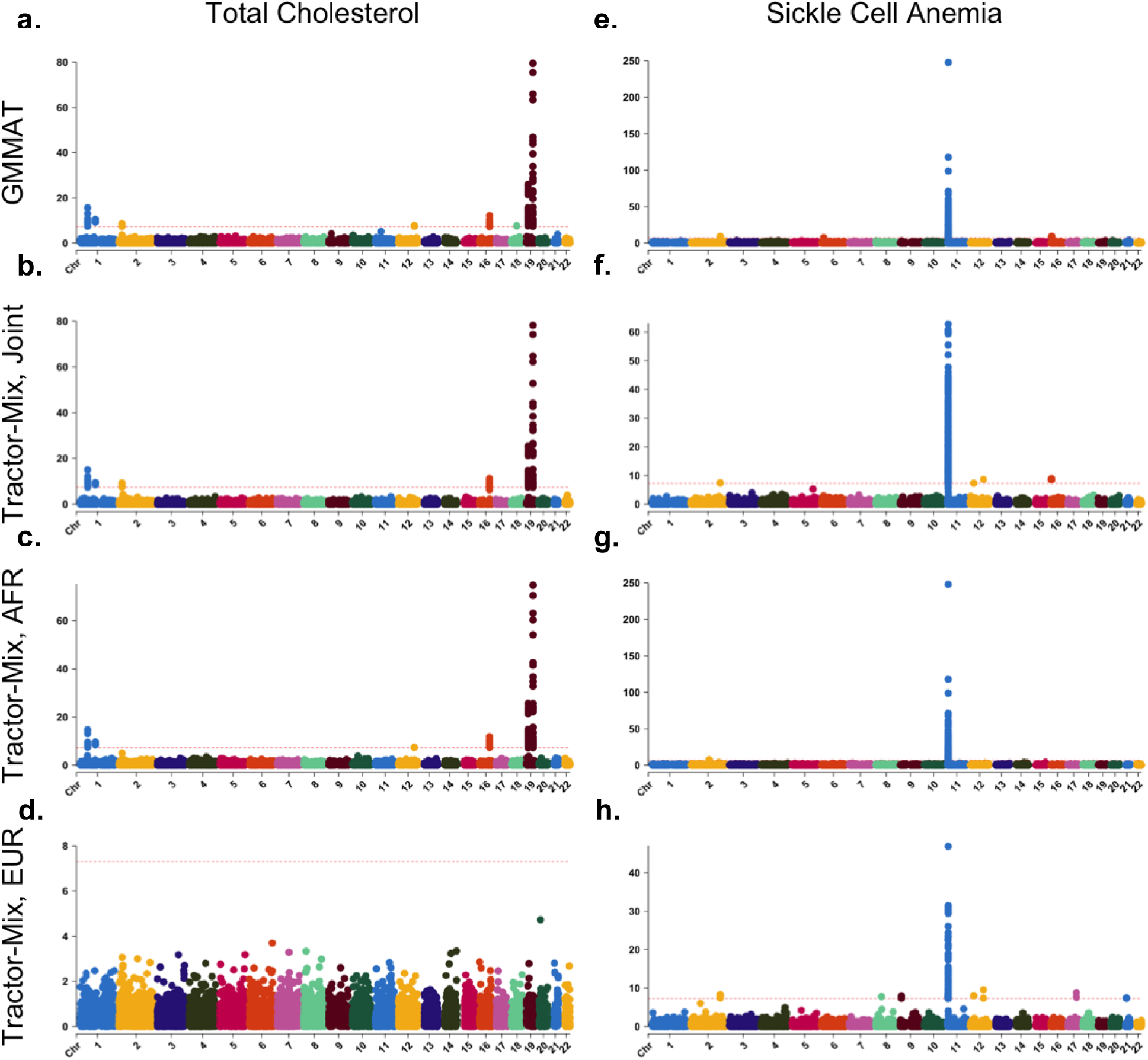
GWAS results from *Tractor-Mix* and *GMMAT* in the UK biobank. We applied GMMAT and *Tractor-Mix* to total cholesterol and si ckle cell anemia in UKBB 2-way AFR EUR admixture. The left columns (a-d) are the Manhattan plots for total cholesterol, and the right columns (e-h) are the Manhattan plots for sickle cell anemia. *Tractor-Mix* identified known hits for both total cholesterol (rs7412) and sickle cell anemia (rs334).

For total cholesterol, *Tractor-Mix* was able to replicate known top hits observed in both GWAS approaches, including identifying the missense variant rs7412 in *APOE* for total cholesterol. This serves as a positive control that our novel 2 d.o.f. model is able to detect real biological signal with comparable power to current gold standard mixed model methods. Compared with the original *Tractor, Tractor-Mix* identified the same lead SN*P* (rs7412) among *APOE* for total cholesterol, but reached a more significant *P* value around 1e-80, compared to 1e-25 for *Tractor*^26^. This is partially due to *Tractor* necessarily removing related samples, resulting in a reduced sample size. We also observe that the risk allele rs7412 exhibits a strong signal in the AFR ancestry background while not in EUR (AFR *P* = 1e-80, EUR *P* not significant). Examining the MAF of this variant as stratified by local ancestry background showed that the risk allele is enriched among AFR ancestries (AFR MAF = 0.120, EUR MAF = 0.068). Examining the *λ*_*GC*_ and QQ plots indicates good control of false positives in these empirical tests, for both the joint *P* value (*λ*_*GC*_ = 0.997) and ancestry-specific *P* values (*λ*_*GC*_ = 1.024 for AFR, *λ*_*GC*_ = 0.964 for EUR). To confirm the results produced by the score test, we extracted the top hits of each chromosome and ran Wald tests on them (Supplementary Table 2), again finding they are largely concordant with the scalable score test approximation (identical if round to 2 significant figures). A comparison of ancestral effect size estimates for total cholesterol top hits can be found in Supplementary Figure 6.

To assess performance in a dichotomous setting, we next examined the summary statistics of sickle cell anemia, a trait with known ancestry-enriched variation. We find that both *GMMAT* and *Tractor-Mix* can replicate the well-known rs334 as the top hit for sickle cell anemia, which encodes a missense variant in the *HBB* gene. rs334 has a large allele frequency divergence across populations (MAF = 0.091 for AFR, 0.0008 for EUR). Additionally, *Tractor-Mix* found several unique variants across the genome not identified by GMMAT, including rs67525852, rs2075208, rs12553528, rs79407438, rs606926, rs12483053, although the biological implications of these variants has not been deeply characterized.

#### Mexico City Prospective Study

We further evaluated the performance of Tractor-Mix in the Mexico City Prospective Study (MCPS) – a highly admixed Latin American cohort of over 150K adults recruited from the Coyoacán and Iztapalapa districts of Mexico City between 1998 and 2004^48^. Extensive family networks are notably embedded within the cohort – 71% of MCPS participants share at least one close relative within the 3^rd^ degree in the study^25^– thereby making MCPS an ideal test case for *Tractor-Mix*. We ran a GWAS of body mass index (BMI), an important risk factor for premature death in Mexico^49^, using three approaches implemented in *PLINK, GMMAT*, and *Tractor-Mix*. To maximize power to detect genetic effects across ancestral backgrounds, we restricted the analysis to MCPS participants with less than 2.5% global ancestry from East Asian and African populations and applied a two-way admixture model including Indigenous American (IAM) and European (EUR) genetic ancestries. Among the resulting set of 33,500 MCPS participants, the mean IAM and EUR ancestry values were 85.1% and 14.9%, respectively (Supplementary Figure 7).

We applied linear models that adjusted for age, sex, and IAM ancestry proportion (to account for population structure) as covariates. All GWAS analyses revealed genome-wide significant associations at loci previously associated with BMI, including the well-known *FTO* locus (Figure 4). Expectedly, the *PLINK* analysis, which did not include any additional adjustment for genetic relatedness, yielded the highest genomic control (GC) lambda inflation factor of 1.40 (Supplementary Figure 8). In contrast, the score test implemented in *GMMAT*, which included a sparse GRM in the fitted null model, resulted in a lower inflation factor of 1.24. *Tractor-Mix* yielded a comparable inflation factor of 1.27 (Supplementary Figure 8). The ancestry-specific *Tractor-Mix* GWAS results for the IAM and EUR ancestries yielded consistent inflation factors of 1.23 and 1.27, respectively.

**Figure 4.**
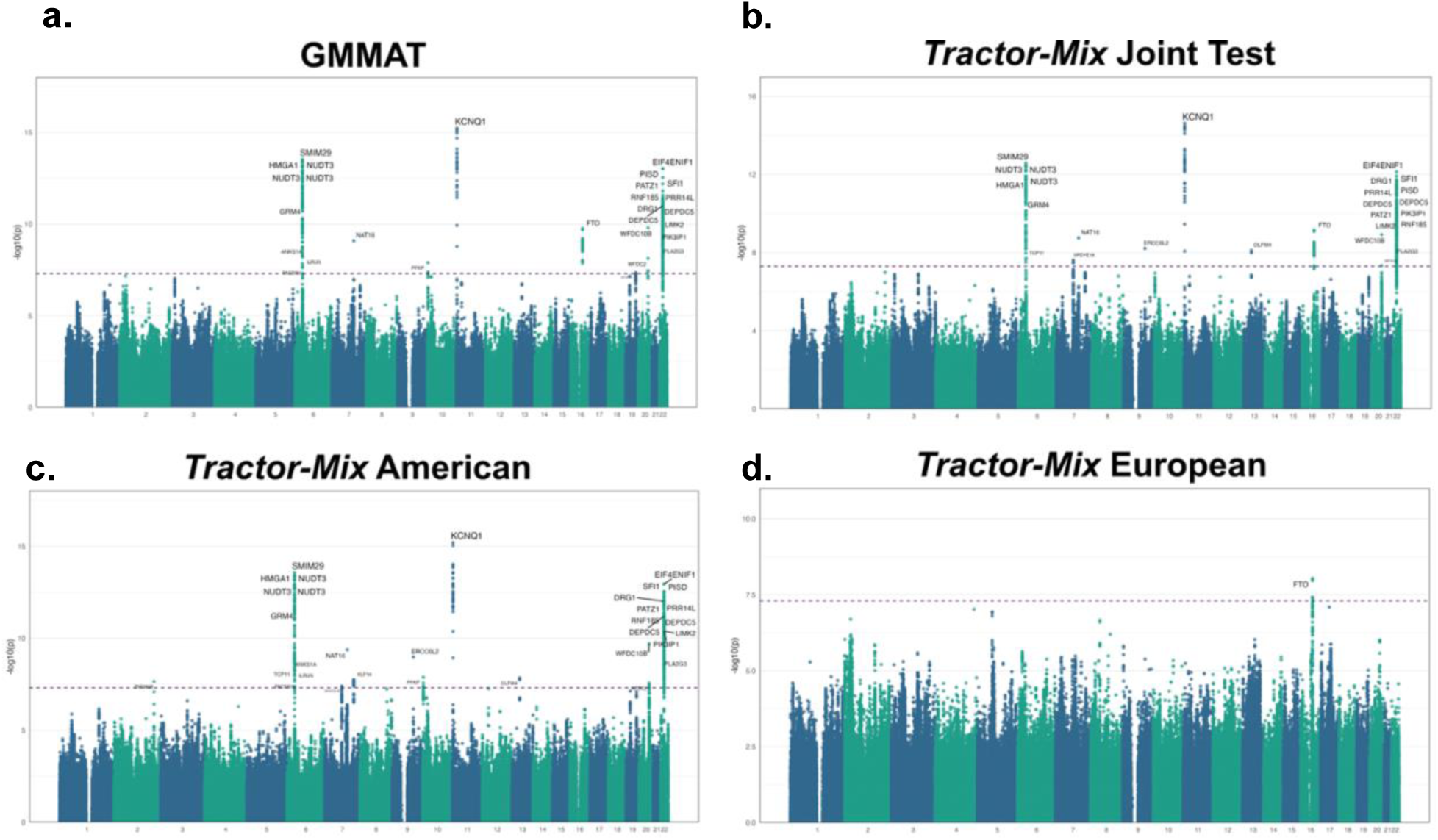
GWAS results for analysis of BMI using GMMAT and Tractor-Mix in the Mexico City Prospective Study. Single-variant association tests were performed on 33,500 MCPS participants with either a score test implemented in GMMAT or a local ancestry-stratified test implemented in Tractor-Mix. A 2-way admixture model allowing for genetic inheritance from inferred IAM and EUR ancestral populations was applied for the Tractor-Mix joint and ancestry-specific tests. Results are shown in Manhattan plots where the nearest genes to the most strongly associated variants at each locus are labeled and the horizontal dashed line indicates the standard genome-wide significance threshold, corresponding to p-value ≤ 5 × 10-8.

Genome-wide association profiles were largely consistent between *GMMAT* and the *Tractor-Mix* joint test (Figure 4, Supplementary Figure 8). However, the ancestry-decomposed association profiles were markedly distinct. Notably, the *FTO* locus, which harbors the strongest genetic association with BMI in Europeans, was found to only be significant in the EUR background among MCPS participants. In fact, this was the only genome-wide significant association in the EUR-specific *Tractor-Mix* analysis (index variant rs11647020, *p-value* = 9.36 × 10^−9^). In contrast, all other genome-wide significant ancestry-decomposed associations were specific to the IAM background, which largely mirrored the *GMMAT* and *Tractor-Mix* joint association profiles (Figure 4). However, most of the genome-wide associations from *Tractor-Mix* IAM analysis mapped within 250kb of known BMI-associated variants that have been reported from studies of predominantly European decent. This suggests that much of the power to detect BMI-associated variants in this analysis involved effect alleles that were segregating appreciably, but not necessarily specifically, among IAM haplotypes that were abundant in the cohort.

Despite the high concordance in *GMMAT* and *Tractor-Mix* GWAS results, the *Tractor-Mix* joint test did detect an association at the *ZNF646PI* locus that did not reach genome-wide significance in the GMMAT analysis (Figure 5, Supplementary Figure 9). Although this variant (rs9568859, *p-value* = 7.67 × 10^−9^) had not previously been associated with BMI *per se*, it has been associated with waist circumference, giving it relevance to the phenotype at hand^50^. Moreover, other SNPs proximal to the *ZNF646PI* pseudogene (located ~92kb from rs9568859), have been previously associated with adiposity traits including BMI and body size^29^. As with other genome-wide significant associations in MCPS, rs9568859 associated with BMI in the IAM but not the EUR background in the ancestry-stratified *Tractor-Mix* analyses (Figure 5). Although the effect allele was common among IAM ancestry (AF = 0.485), it was also present in European (0.113) and African ancestries (0.239). This indicates that *Tractor-Mix* can identify novel trait associations and yield ancestry-specific marginal effect sizes, even when causal variants are likely to be shared across populations. Overall, these results corroborate that *Tractor-Mix* can disentangle genetic associations by ancestry while adjusting for genetic relatedness in a highly admixed and extensively related Latin American population.

**Figure 5.**
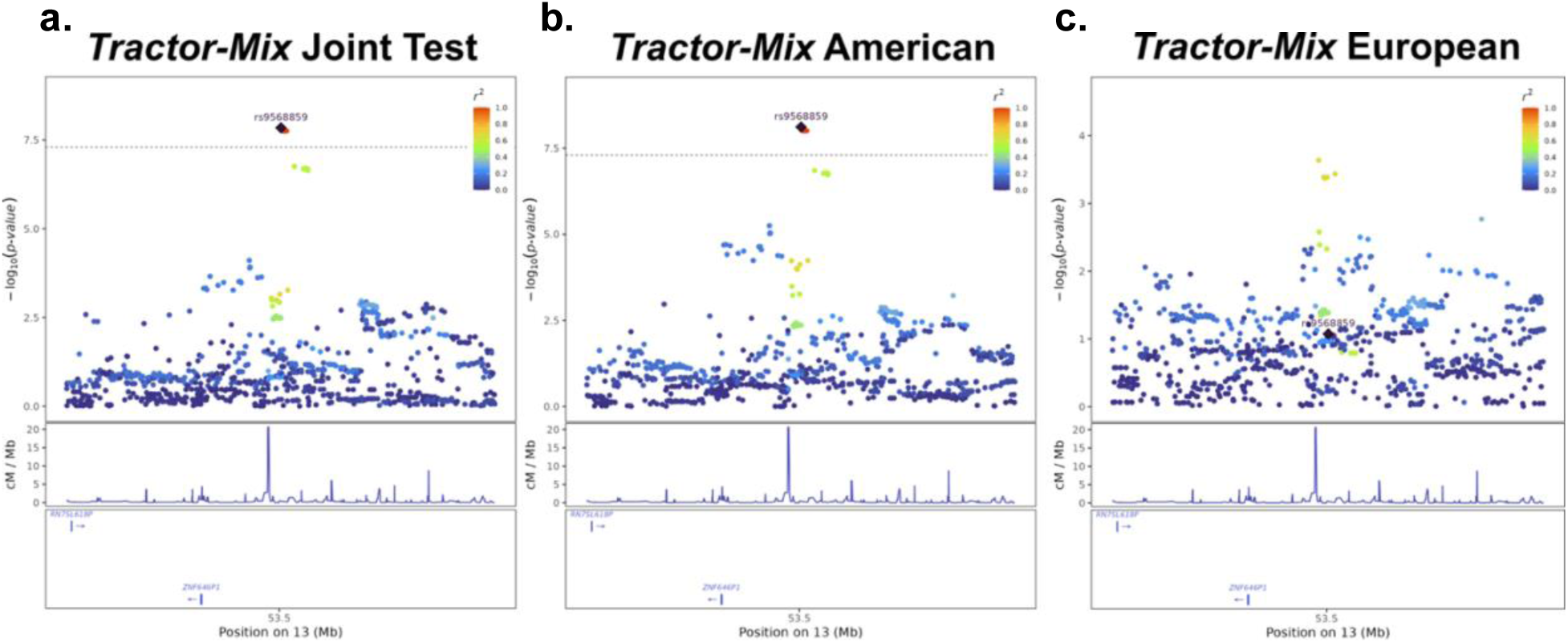
Locus plots at the *ZNF646P1* locus from the GWAS of BMI in the MCPS cohort. *Tractor-Mix* associations for variants mapping within 250kb of the index variant rs9568859 are shown with color corresponding to pairwise LD *r*^*2*^ with respect to this SNP. rs9568859 is an intergenic variant that is located ~92kb upstream of the TSS of *ZNF646P1*, which encodes Zinc Finger Protein 646 Pseudogene 1.

#### Yale-Penn cohort

We additionally applied *Tractor-Mix* on the Yale-Penn Cohort to study the genetic basis of alcohol related traits. Our results suggest ancestry-specific associations for AUD in AFR ancestry (*Supplementary Information*, Supplementary Figure 10). Importantly, this additional validation highlights the novel model presented here is well controlled for Type I error for highly polygenic traits.

## Discussion

Here, we describe a local ancestry informed GWAS framework that can handle admixed cohorts with high degrees of relatedness, filling a significant gap in the existing methodological space. Leveraging local ancestry deconvolution in GWAS allows for the generation of ancestry-specific estimates (*P* values, effect sizes) for risk alleles that can empower downstream efforts, such as fine-mapping and PRS^27^. It further enables localization of GWAS signal onto specific ancestry backgrounds, permitting identification of loci uniquely discoverable in, or variants enriched in, given ancestry backgrounds. This can provide meaningful context relevant for variants that may inform personalized medicine.

In designing novel methods, there is always a choice for which area of parameter space the model should be optimized. We have chosen to optimize our model in the service of identifying ancestry-enriched loci, gaining power over traditional methods from ancestral heterogeneity. This necessarily comes at a trade-off of a reduction in power to identify loci with shared effects across populations. The rationale behind our choice is that, given the massive sample sizes and extensive prior work on European datasets (~80% of all GWAS are conducted on European samples^6,9^), such ancestry-enriched loci are more likely to be novel, and thereby provide value beyond those shared loci that are likely to be already represented in existing large GWAS studies. Importantly, the power loss in identification of shared loci is minimal enough that in most cases such loci will still pass genome-wide significance (Figure 2). We thus have optimized our tool to identify novel biology in less well studied backgrounds as the priority.

Further unpacking heterogeneity, effect size heterogeneity can be categorized into either causal (the effect of the functionally impacting variant is truly different depending on the ancestral haplotype on which it falls) or marginal (the effect at the common SNPs tagging causal loci is different due to ancestral differences). While causal effects are generally assumed to be similar across ancestries^51–55^, (though there are some demonstrations of causal heterogeneity across different ancestries^56–59^), tagging differences manifest as differences in estimated effect sizes across ancestry backgrounds, inducing effect size heterogeneity even when there is a shared causal variant with the same effect^1,37,42,51,60,61^. We underscore that *Tractor-Mix* will benefit from heterogeneity both in true causal effects as well as via marginal, tagging impacts.

Besides the capability of controlling for family structure, one notable difference between *Tractor-Mix* and *Tractor* is that *Tractor-Mix* does not condition on overall local ancestry at a site (i.e. the ‘hapcounts’ term in the original *Tractor* model). Recent papers have explored the role of conditioning on local ancestry for GWAS analysis in admixed populations^36,61,62^. Briefly, conditioning on local ancestry can absorb the linkage disequilibrium signal induced by admixture events (formally called Admixture LD). Therefore, the ancestry-specific summary statistics obtained from *Tractor* have intuitive interpretations: they are equivalent to marginal effect size estimates obtained from homogeneous populations. Conditioning on local ancestry has the additional benefit of narrowing GWAS peaks to smaller regions. For these reasons, the original *Tractor* methodology elected to include this term. However, conditioning on local ancestry can reduce statistical power due to collinearity between that term and ancestry-specific allele counts. Therefore, we view overall haplotype-level local ancestry as an optional term in GWAS analysis – whether to include this term is a tradeoff between effect size interpretability and statistical discovery power. In *Tractor-Mix*, we chose to exclude the local ancestry term primarily to relieve computational burden. As *Tractor-Mix* employs a score test framework, including the local ancestry term would require fitting null models for every local ancestry segment. This can result in fitting hundreds to thousands of null models, which is computationally expensive. Dropping the local ancestry term, as has been suggested in some prior work^63^, addresses this challenge, and additionally empowers identification of ancestry-specific loci. We feature here a GWAS of sickle cell anemia (Figure 3), a trait with well characterized ancestry background-enriched signals^64,65^, to highlight the utility of *Tractor-Mix* in this use case.

There are some important limitations of *Tractor-Mix* to consider. Firstly, we performed systematic evaluations of *Tractor-Mix* on two-way admixture, but three-way admixture (or higher) has not been examined in this study. With multi-way admixture, *Tractor-Mix* might not achieve similar performance as two-way admixture, as more free parameters are introduced in the model, resulting in a higher statistical penalty. Deconvolving risk alleles to more local ancestries can also introduce sparsity of the genotypes. In general, we therefore advise limiting the number of component ancestries modeled to just those with sizeable contributions (e.g. >=10% average global ancestry) in your dataset to maximize power, and advise care when modeling multi-way admixed populations.

Secondly, we implement *Tractor-Mix* in R with large scale matrix multiplication. Although our inclusion of a sparse GRM implementation, combined with our multi-core parallelization, significantly improves the computational speed, *Tractor-Mix* is likely to be slow in large-scale settings (e.g. >=tens of thousands of samples and tens of millions of markers).

Finally, *Tractor-Mix* inherits functions from *GMMAT*; it is thus similarly robust for most case-control studies but might produce biased results when the case-control ratio is highly imbalanced, or when ancestry-specific allele counts are low. To address this, we propose dropping ancestry-specific genotype dosage terms that have low AC and computing marginal *P* values and effect sizes, as described in the *Methods*. This approach effectively reduces false positives and allows for ancestry-specific summary statistic generation in the background harboring the signal, however the output would have a different interpretation from that of the joint model. We track the model utilized for each variant should users wish to filter to only variants experiencing the full model. To effectively address the challenge of modeling variants with low ancestry-specific AC, more advanced techniques, such as saddle point approximation, could be implemented in the future^29^.

In sum, *Tractor-Mix* offers a powerful and reliable methodology for association analyses of complex traits in admixed populations with relatedness. *Tractor-Mix* not only retains the strengths of the original *Tractor* GWAS to powerfully identify ancestry-specific or enriched signals, but also offers further benefits in the form of boosted statistical discovery power, and highly effectively controlling for false positives via a GRM. By effectively accounting for population stratification and relatedness in the context of admixed populations, our approach addresses longstanding challenges in genomic research, paving the way for more accurate and representative genetic discoveries. This is particularly vital as biobanks and large-scale genetic data collections are increasingly encompassing more diverse and complex populations^34,66,67^, and the population as a whole is becoming increasingly admixed over time^8,68^. We hope that our work facilitates the further study of admixed populations in GWAS and thereby accelerates insights into the genetic underpinnings of important phenotypes across populations.

## Methods

### Generalized linear mixed model for jointly testing ancestry-specific markers

Similar to the original *Tractor*, we jointly model ancestry-specific markers with local ancestry inference (see the *Tractor*^26^ paper for a detailed discussion), while additionally taking sample-relatedness into account under the mixed effect model framework. Without a loss of generality, we illustrate *Tractor-Mix* for 2-way admixed cohorts:

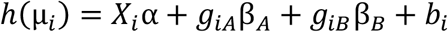

where *i* is the index for individuals, *h*(μ_*i*_) = μ_*i*_ for continuous traits, and *h*(μ_*i*_) = *logit*(μ_*i*_) for binary traits. *X*_*i*_ is a vector of covariates (e.g. intercept, age, sex, PCs), where PCs can be obtained from a software such as PC-AiR^31^; *α* is a vector of effect sizes for covariates. *g*_*iA*_, *g*_*iB*_ are ancestry-specific markers partitioned by local ancestry background (e.g. *g*_*iA*_ is the ancestry-specific genotype dosage for ancestry A, *g*_*iB*_ is the ancestry-specific genotype dosage for ancestry B). *b* is the random effect term that is distributed as MVN(0, *τ*^2^Φ), where Φ is a genetic relationship matrix (GRM) or kinship matrix, and *τ*^2^ is the additive genetic variance.

To test the hypothesis *H*_0_: β_*A*_ = β_*B*_ = 0, we used a 2 degree of freedom Rao’s score test to obtain a joint *P* value. We leverage the package *GMMAT* to fit the null model *logit*(μ_*i*0_) = *X*_*i*_*α* + *b*_*i*_ and to obtain 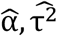 with a penalized quasi-likelihood approach (see Chen *et al*. 2016 for a detailed discussion)^28^. The score for *H*_0_ is given by 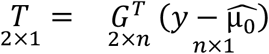 where 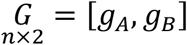 is a n × 2 matrix (stacking ancestry-specific genotype dosage), 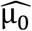 is the fitted value under *H*_0_ (shared across all variants). To get the covariance matrix for the score, we have 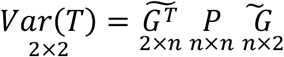 where 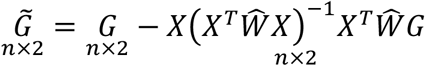 and 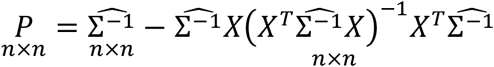. Notice that 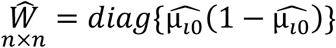 and 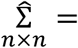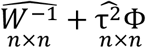. The joint test statistics for *H*_0_: β_*A*_ = β_*B*_ = 0 can be obtained by 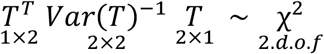.

Rao’s score test does not directly fit the full model, but the ancestry-specific effect sizes can be approximated by 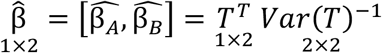. We can also estimate ancestry-specific standard errors with 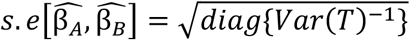, which allows us to approximate *P* values for β_*A*_, β_*B*_ without fitting the full model. We note that the standard error approximation is valid under the null hypothesis, but can deviate from truth under the alternative hypothesis. Although less scalable, we implemented a 1 d.o.f. Wald test by directly fitting the full model by adapting the *GMMAT*^28^ source code. This method allows us to obtain ancestry-specific *P* values, given as 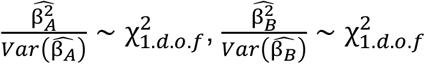.

As allocating risk alleles to local ancestry tracts can reduce the allele counts, we were concerned that *Tractor-Mix* might have increased false positive rates for dichotomous traits. One solution for this is to incorporate the saddle point approximation (SPA) strategy, similar to FastSPA^69^ and SAIGE^29^. However, extending SPA to high dimensional genotype dosage space is challenging and therefore a direction of future work. Here, we instead implemented a simpler strategy: we let the user define a minimum allele count threshold (default is 50). We then conduct hypothesis testing with ancestry-specific genotype dosages that pass this threshold. A joint *P* value will be calculated if all the modeled ancestries pass this threshold, otherwise ancestries failing the AC cutoff will be dropped from the model and only the marginal *P* values will be generated. Although we will thereby not estimate the joint effect size of all ancestries for all variants, this strategy allows us to reliably estimate the marginal effects of the ancestries harboring the GWAS signal.

### Simulations to assess the False Positive rate

To evaluate the false positive rate, we simulated 2-way admixed AFR-EUR individuals with a high degree of relatedness. We simulated a cohort with 2,500 independent and 2,500 related samples (from 250 families, Supplementary Figure 1), with 1,000,000 variants for each person. For independent samples, their global ancestry proportion (measured with AFR being the index ancestry) were generated either from a beta distribution Beta(8,12) or Beta(12,8) with 50% of the chance. For family data, we drew global ancestry proportions from either Beta(8,12) or Beta(12,8) with 50% of the chance for all founders of the family (founders in the same family have the same global ancestry proportion). Offspring genotypes/local ancestry are then randomly inherited from their parents using the gene dropping^70^ method. This simulation strategy (including the pedigree) is adapted from the PC-AiR paper^31^, and is intended to mimic assortative mating.

In this simulation, we incorporated local ancestry for every independent individual and founder, with lo*c*_*ijA* ∈{0,1,2}_ ~ *B*inom(2, *Adm*_*iA*_) and lo*c*_*ijE* ∈{0,1,2}_ = 2 − la*c*_*ijA*_. Here, A*dm*_*iA*_ is the AFR global ancestry proportion for an individual, and la*c*_*ijA*_ is the AFR local ancestry for individual i at locus j. Ancestry-specific markers for independent individuals and founders are then drawn from a binomial distribution with *g*_*ijA* ∈{0,1,2}_ ~ *B*inom(*lac*_*ijA*_, *AF*_*jA*_), *g*_*ijE* ∈{0,1,2}_ ~ *B*inom(*lac*_*ijE*_, *AF*_*jE*_), where A*F*_*jA*_, A*F*_*jE*_ represents ancestry-specific allele frequencies (AFR, EUR) for variant j. We note that la*c*_*ijA*_ + la*c*_*ijE*_ = 2, and *g*_*ijA*_ + *g*_*ijE*_ = *g*_*ijGWAS*_. We also note that A*F*_*jA*_, A*F*_*jE*_ are generated from the Balding-Nicoles model^71^ parameterized by *F*_*ST*_ = 0.1, with the baseline allele frequency randomly drawn from a uniform distribution AF ~ *U*nif(0.1,0.9).

Once we obtained the ancestry-specific genotype matrices, we simulated the phenotype under the null model to examine the rate of type 1 error with:

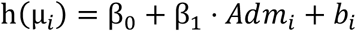

where *Adm*_*i*_ is the global admixture proportion, and is intended to simulate population stratification. *b*_*i*_ is the random effect generated from MVN(0, *τ*^2^Φ). β_1_ is the effect size for the global admixture proportion and β_0_ is the intercept term (for dichotomous phenotypes, this term is adjusted to meet the desired prevalence). Φ is the true kinship matrix derived from the pedigree (Fig S1). During the simulation, β_0_ is fixed at 1. β_1_ was set to either of [0.5, 2]; the variance component parameter *τ*^2^ was set to either of [1,2,5].

In realistic datasets, population structure and family structure are often unknown, and need to be estimated using genotype information. In other words, we need to estimate *Adm*_*i*_ and Φ. Here, we explored different combinations of *Adm*_*i*_ and Φ estimators to ensure *Tractor-Mix* consistently has a well-controlled false positive rate (see the 8 models described in Fig 1).

For the fixed term: Standard PCs were computed using all samples without removing related samples. PC-AiR first partitions samples into independent samples and related samples. Then it constructs the PC space only with independent samples, and projects related samples back into that PC space^31^. Theoretically, the PC-AiR method should thereby better capture population structure. The admixture proportion is the true fixed term, and served as a ground truth in our simulation.

For the random term, the GRM refers to the standard estimator of the coefficient of relationship, computed by 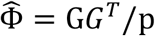, where G is the standardized genotype matrix (mean = 0, var = 1), and p is the number of markers used. PC-Relate refers to the method developed by Conomos *et al*., where family-relatedness is calculated by removing population-level relatedness^32^. Theoretically, PC-Relate should resemble true kinship matrix. The kinship estimator was obtained from the pedigree and served as ground truth in our simulation. Both PC-AiR and PC-Relate are calculated using *GENESIS*^*33*^.

We ran varying numbers of simulation iterations, affording us the ability to assess two significance levels, *α* = 5 × 10^−2^, *α* = 5 × 10^−4^ as the empirical type 1 error rates. We then compared false positives for the 8 different models under different parameter settings (varying β_1_, *τ*^2^). We did not use the conventional *α* = 5 × 10^−8^ as our threshold in this benchmark, as for each model, billions of statistical tests would be required to accurately evaluate false positives under this *α*, making feasibility intractable. However, we conducted one larger-scale false positive evaluation for *Tractor-Mix* with true covariates and GRM (Admixture proportion + Kinship). With ~120M *P* values for continuous phenotypes, and ~22M *P* values for dichotomous phenotypes, we find we were able to accurately evaluate FP for *α* = 5 × 10^−6^ (Supplementary Table 1).

### Simulations for discovery power and effect size estimation

We performed numerical simulations to test the empirical power of *Tractor-Mix* for GWAS in an admixed cohort with relatedness. In the cohort, we have 1,000 independent samples and 1,000 relatives (see Supplementary Figure 1a for the pedigree). Each individual has 80% global AFR ancestry in expectation. For simplicity, we fixed the MAF for both AFR and EUR ancestry components: *AF*_*A*_ = 0.2; *AF*_*E*_ = 0.3. We generated both dichotomous and continuous monogenic phenotypes according to:

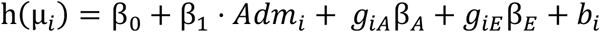

We note that for dichotomous phenotypes, β_0_ is chosen to control for overall prevalence, which was set to be 20%.

To examine how effect size heterogeneity across local ancestry tracts can affect the statistical power for *Tractor-Mix*, we varied the ratio of β_*A*_: β_*E*_ in each simulation. We tested scenarios where effect size heterogeneity is large (1:0 and 0:1), moderate (1:0.5) and nonexistent (1:1). While examining the statistical power of *GMMAT* vs *Tractor-Mix*, we also obtained effect size estimates from *Tractor-Mix* via the score test (approximated by 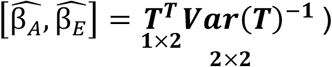 or Wald test (directly fitting the full model). 100 trials were run for each simulation setting.

### Simulations evaluating ancestry-specific *P* value approximation

Similar to our power simulations, we examined the concordance of *P* values produced by the score test and the Wald test. We simulated both continuous and dichotomous phenotypes for a cohort with 1,000 admixed samples (500 independent, 500 related; Supplementary Figure 1).

For continuous traits, we set *τ*^2^ = 1, *σ*^2^ = 1 as the default. Under the null hypothesis, we set [β_0_, β_1_, β_*A*_, β_*E*_] = [1, 2, 0, 0]; under the alternative hypothesis, we set [β_0_, β_1_, β_*A*_, β_*E*_] = [1, 2, 1, 0.5]. For dichotomous traits, we set *τ*^2^ = 1 as the default. Under the null hypothesis, we set [β_1_, β_*A*_, β_*E*_] = [2, 0, 0]; under the alternative hypothesis, we set [β_1_, β_*A*_, β_*E*_] = [2, 2, 1]. We varied the intercept to accommodate the desired prevalence of 0.2, without consideration of the kinship matrix b.

### Empirical testing on the UK Biobank

We processed 9,220 2-way AFR-EUR admixture with approximately 8.9 M markers in UKBB (hg19 build). The variants were QC-ed such to contain only biallelic SNPs with > 0.5% MAF, > 95% call rate, *P* value for HWE less than 1 × 10^−6^, and INFO scores > 0.8.

For construction of the PCs and GRM, we performed LD pruning with PLINK2^72^, with the ‘--indep-pairwise 500kb 0.2’ flag. King-Robust was used to infer relatedness. We extracted continental AFR and EUR individuals from the Thousand Genomes Project reference data and merged these with the pruned UKBB data, resulting in a combined dataset with 32,132 independent variants for all 10,384 samples (9,220 UKBB + 1,164 TGP). Specifying *k*=2, we then ran ADMIXTURE on this merged dataset to confirm samples were indeed two-way AFR-EUR admixed (Supplementary Figure 4). The global ancestry was visualized with ‘popkin’ package in R.

We used *Shapeit5* with default parameters for statistical phasing^73^, providing the 1,164 continental AFR and EUR from the Thousand Genomes Project as the reference panel^74^. Using the same reference panel, we used *RFMix2* for local ancestry inference^75^. The genetic map files used in Shapeit5 and RFmix2 (b37) were obtained from the SHAPEIT github^76^.

*Tractor-Mix* analysis requires 1) estimated global ancestry or PCs, 2) estimated GRM, 3) partitioned genotype files, and 4) phenotype and covariate files (Extended Data Figure 1). For 1) and 2), we applied PC-AiR and PC-Relate on our UKBB dataset according to the GENESIS vignette^33^. The KING GRM estimated with PLINK2 was used in PC-AiR. Top 2 principal components from PC-AiR were used for computation with PC-Relate. The estimated GRM from PC-Relate was rescaled (X2 to make the diagonal elements close to 1) and masked such that elements less than 0.05 were set to 0. The masking procedure converts a full GRM to a sparse GRM, which increases computational efficiency and possibly reduce false positives in GWAS. To obtain 3) partitioned genotype files, we applied the extract_tracts.py step from *Tractor*. The relevant outputs are ‘anc0.dosage.txt’ and ‘anc1.dosage.txt’, which record risk alleles allocated to the two component ancestries. To obtain 4) phenotype and covariate files, we downloaded UKBB data and used total cholesterol (30690), LDL cholesterol (30780) and sickle cell anemia (282.5) as our test traits. The null model was fitted with the ‘glmmkin()’ function from *GMMAT*, providing age, sex, and the top 5 PCs from PC-AiR as covariates, and using the sparse PC-Relate as the estimated GRM. After intersecting the phenotype files and genotype files, the total sample sizes for total cholesterol, LDL, and sickle cell anemia are 8,583, 8,566, and 8,378, respectively. To compute the 1 d.o.f. *GMMAT* test, we applied a standard score test (‘glmm.score()’) on PLINK format data, with genotypes specified to be non-centered (‘center = F’). To compute the 2 d.o.f. *Tractor-Mix* test, we applied ‘TractorMix.score()’. The output QQ plot and Manhattan plot are visualized with the CMplot() R package. When applying *Tractor-Mix* on sickle cell anemia, we used the default AC = 50 as the threshold for binary traits. Joint models were used for variants with both ancestry-specific genotype dosages above this threshold. Otherwise, the ancestry-specific genotype dosage with low AC was first dropped, and marginal models were applied.

### Empirical testing of the Mexico City Prospective Study

Autosomal genotypes assayed on the Illumina Global Screening Array (v2) and ADMIXTURE-based ancestry proportion estimates were obtained for participants from the Mexico City Prospective Study (MCPS) – a prospective study of over 150K adults recruited from the Coyoacán and Iztapalapa districts of Mexico City between 1998 and 2004^25,48^. To ensure a two-way admixture model, we excluded individuals with greater than 2.5% ancestry from either inferred African or East Asian ancestral populations, retaining only those with admixture predominantly from Indigenous American (IAM) and European (EUR) ancestries. This resulted in a set of 33,500 participants selected for GWAS analyses of BMI. The mean IAM and EUR ancestry values in this subset were 85.1% and 14.9%, respectively. Given the high prevalence of diabetes in MCPS which has been found to be inversely-associated with BMI in this cohort^49^, individuals with a self-reported previous-diagnosis of diabetes or glycated hemoglobin (HbA1C) levels ≥6% were excluded.

GWAS analyses were performed using Trans-Omics for Precision Medicine (TOPMed)-imputed genotypes (obtained via the TOPMed Imputation Server) with imputation INFO score > 0.8. Variants were further restricted to biallelic SNPs with MAF ≥0.5%, genotype missingness <5%, and not significantly deviating from Hardy-Weinberg equilibrium (at α = 1 × 10^−6^). A total of 8,051,365 autosomal SNPs met these criteria. Variant positions corresponded to reference genome GRCh38 (hg38). To facilitate local ancestry inference, MCPS genotypes meeting quality control thresholds were merged with variant calls from whole genome-sequenced samples from the 1000 Genomes Project ([TGP]; phase 3) and Human Genome Diversity Project (HGDP). Following allele harmonization and removal of mismatched and ambiguous variants, a total of 5,887,724 biallelic SNPs were retained in the merged dataset.

Genotypes from the merged MCPS and reference dataset were phased with SHAPEIT v5.1.1. using default parameters and local ancestry inference (LAI) was performed with RFMix v2. For LAI, a two-way admixture model was applied using 408 IAM and 658 EUR reference samples from the TGP and HGDP datasets. Specified parameters used in RFMix included 15 generations since admixture, 5 terminal nodes for the random forest classifier, and 5 rounds of the expectation maximization algorithm for the ‘re-analyze reference’ option.

We developed a custom pipeline to manage memory constraints and parallelize computations to facilitate ancestry estimation in large-scale data (i.e. >10K samples). This entailed splitting per-chromosome genotype data into chunks of sample subsets that were analyzed in parallel with RFMix. Following LAI analysis, results from all chunks were merged into a single dataset for each chromosome. To ensure consistency across chunk boundaries, a complete set of local ancestry segments were delineated using the *disjoin* function from the GRanges package in R.

The *extract tracts* program from Tractor was then used to deconvolute genetic markers by local ancestry. Population structure was characterized using PC-AiR and involved LD pruning with an LD r^2^ threshold of 0.1 to select independent SNPs. The PC-AiR algorithm requires pairwise kinship coefficients, computed with the KING-robust method. Due to computational limitations in handling the kinship matrix in R, a custom script was developed to create a sparse matrix representation of kinship. PC-Relate was subsequently applied to estimate recent genetic relatedness and utilized principal components from PC-AiR to account for more distal population structure.

The genetic relationship matrix (GRM) estimated from PC-Relate was subsequently used to fit a null linear mixed model for BMI, taking into account sex, age, and IAM ancestry proportions as fixed effect terms, and GRM as the random effect term.

GWAS analyses of BMI were performed using PLINK v2, GMMAT, and *Tractor-Mix* with sex, age, and IAM ancestry proportions used as covariates. A standard linear regression model was implemented in PLINK to test for association between genotype and BMI. For GMMAT, the ‘glmm.score()’ function was used to implement a score test on genotype data in PLINK format with non-centered genotypes (‘center = FALSE’). This analysis incorporated the null model described above, with the aim of enhancing statistical power and precision by explicitly accounting for pairwise relationships in the GRM.

*Tractor-Mix* was run using the same null model for BMI as used in the GMMAT analysis and ancestry-specific genotype dosages as independent variables. Due to differentiated and/or low allele counts (AC) when stratifying by genetic ancestry, we applied strict AC filtering. Variants were retained in the full ancestry model only if the observed AC exceeded 50 in each ancestry group (i.e. IAM and EUR), otherwise variants not meeting this threshold were excluded from the full ancestry test and ancestry-specific effects were not estimated. However, such variants may still have been included in the joint test implemented in Tractor-Mix if AC > 50 in the full dataset (i.e. not accounting for the specific ancestral background of each allele). This approach minimizes type I errors while preserving statistical power when allele counts are sufficient across ancestries. Ancestry-specific effect sizes, standard errors and *p*-values, and joint *p*-values estimated with Tractor-Mix were compared with summary statistics from the PLINK and GMMAT analyses. Potentially novel BMI-associated loci were implicated by assessing if genome-wide significant variants mapped within 250kb of any previously-reported variants associated with BMI in the NHGRI-EBI GWAS Catalog (accessed February 20th, 2025). Variants were further annotated for prior trait associations using the GWAS Catalog, genomic and functional annotations using the Ensembl Variant Effect Predictor, and ancestry-specific allele frequencies using the MCPS Variant Browser.

### Software implementation

*Tractor-Mix* leverages a similar framework to *Tractor*, including some shared preliminary steps: 1) Phasing and local ancestry inference; 2) Optionally recovering phasing errors; 3) Extracting tracts and computing local ancestry and ancestral genotype dosages. These steps have been described in detail in the original *Tractor* manuscript ^26^, and are inherited in *Tractor-Mix*. Once we obtain ancestry-specific risk alleles count, we can use *Tractor-Mix* to compute the *P* values using Rao’s score test. Rao’s score test includes two steps: 1. Fit the null model; 2. Scan over all genetic markers to compute the *P* values. For step 1, *Tractor-Mix* share the same code as *GMMAT*. For step 2, we developed R code for both dense GRM and sparse GRM (with the R package ‘*Matrix*’). We enabled multi-core calculation with R packages ‘*doParallel*’ and ‘*foreach*’. Although the R implementation is not optimized for computational speed, a moderately sized GWAS analysis can be done within a reasonable amount of time. In our UKBB analysis with 9,000 samples and 8.5M variants, it took approximately 10 hours per trait with 22 chromosomes run in parallel on a typical high performance computing cluster setup (using sparse GRM, 32GB RAM, 8 CPUs per chromosomes).

## Data Availability

All data produced in the present study are available upon reasonable request to the authors

## Data availability

Summary statistics from all empirical runs will be deposited into the GWAS catalog for open access.

## Code availability

Code to run the Tractor-Mix model and reproduce the empirical analysis presented here are available on GitHub at, respectively: https://github.com/Atkinson-Lab/Tractor-Mix-manuscript and https://github.com/Atkinson-Lab/Tractor-Mix.

The software utilized in this project included:

Shapeit5: https://github.com/odelaneau/shapeit5

RFMix2: https://github.com/slowkoni/rfmix

Tractor: https://github.com/Atkinson-Lab/Tractor

GMMAT: https://github.com/hanchenphd/GMMAT

GENESIS: https://github.com/UW-GAC/GENESIS?tab=readme-ov-file

Plink2: https://www.cog-genomics.org/plink/2.0/

Simulation & analysis: https://github.com/Atkinson-Lab/Tractor-Mix-manuscript

Tractor-Mix: https://github.com/Atkinson-Lab/Tractor-Mix

## Acknowledgements

We thank Patrick Turley for helpful discussions in the formulation of this methodology. This research has been conducted using the UK Biobank Resource under Application Number 95179. We thank the MCPS study participants and staff. The MCPS is a long-standing scientific collaboration between researchers at the National Autonomous University of Mexico and the University of Oxford and has received funding from the Mexican Health Ministry; the National Council of Science and Technology for Mexico; the Wellcome Trust [058299/Z/99]; Cancer Research UK; the British Heart Foundation [RE/13/1/30181]; Kidney Research UK [MR/R007764/1] and the UK Medical Research Council [MC_UU_00017/2, MR/Z504543/1].

## Author Contributions

E.G.A. designed and supervised the project. T.T. constructed the model and conducted simulation analyses and empirical analysis of the UKB. A.V. and J.M. conducted empirical analysis of the MCPS and Yale-Penn cohort, respectively. N.S. advised on code construction. J.B., J.A., P.K., R.T., J.G. and H.K. contributed data. J.T. and J.M. supervised empirical analyses. W.Z. and K.Y. advised on model creation. T.T. and E.G.A wrote the manuscript, with contributions from A.V. and J.T.. All authors read and provided feedback on the manuscript.

## Rights retention

For the purpose of Open Access, the author has applied for a CC BY public copyright licence to any Author Accepted Manuscript (AAM) version arising from this submission.

## Ethics declaration

The authors declare no competing interests.

## Data Availability

Data from the Mexico City Prospective Study are available to bona fide researchers. The study’s Data and Sample Sharing policy (https://www.ctsu.ox.ac.uk/research/mcps) can be viewed (in English or Spanish). The questions utilized in the study, along with the available study data, can be accessed and reviewed through the study’s Data Showcase (https://datashare.ndph.ox.ac.uk/mexico/).

## Extended Data

**Extended Data Figure 1.**
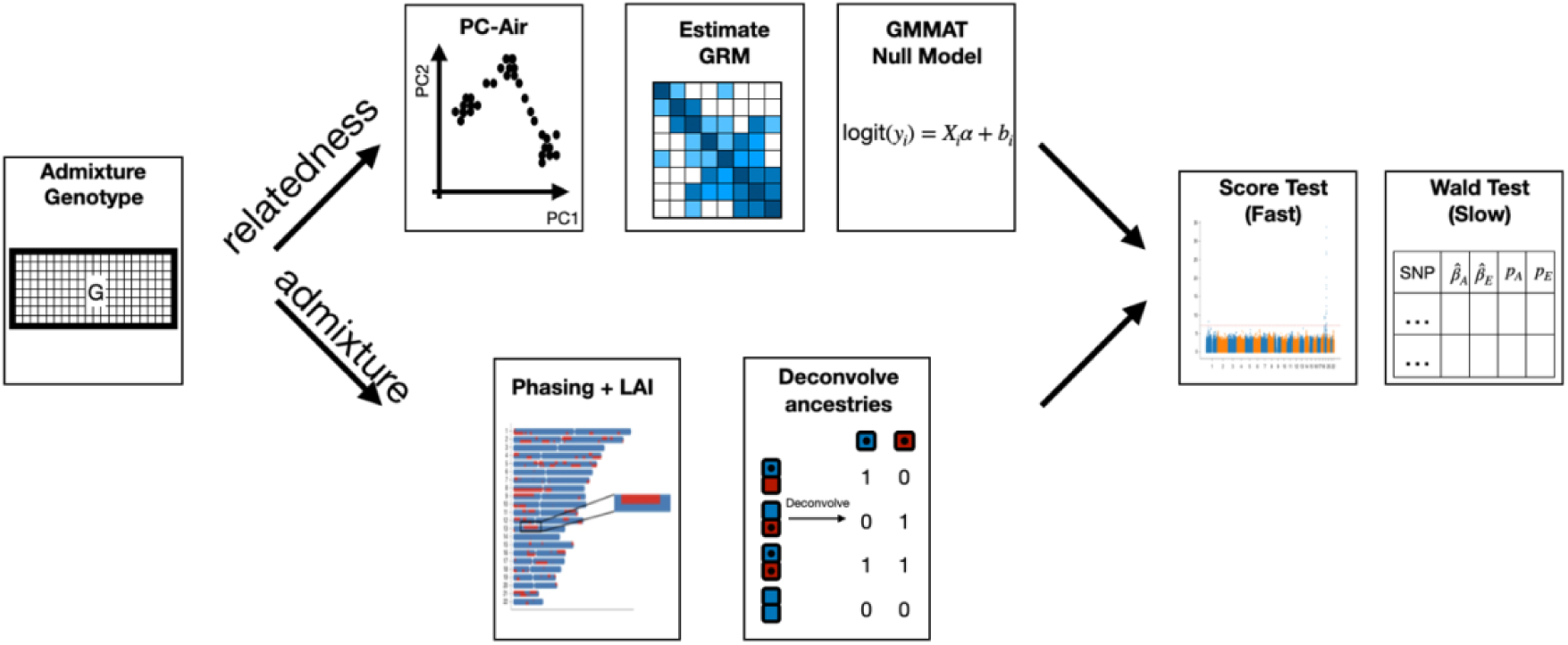
The *Tractor-Mix* pipeline. *Tractor-Mix* consists of 6 steps: 1. Phasing (*Shapeit5*) and local ancestry inference (*RFMix2*) 2. Local ancestry dosage computation with *Tractor* (ExtractTracts.py) 3. PC-AiR with *GENESIS* (or other global ancestry estimators) 4. PC-relate with GENESIS (or other GRM estimators) 5. Fit the null model with GMMAT 6. Calculate joint *P* values, alongside ancestry-specific effect sizes and *P* values with *Tractor-Mix*

**Extended Data Figure 2.**
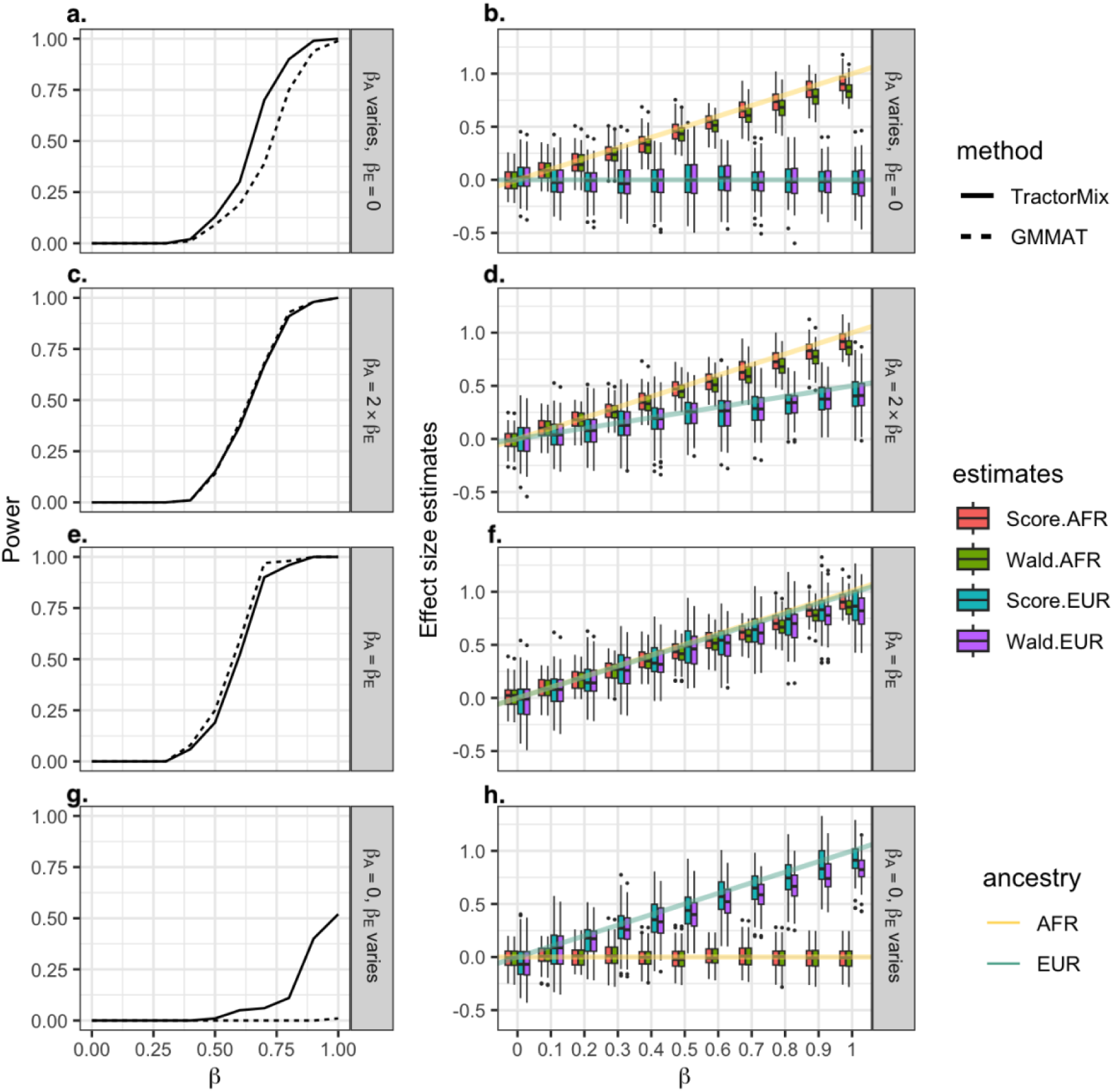
Power and effect size estimates for dichotomous traits. Similar to the continuous traits, we evaluated the power and effect size estimates for dichotomous traits. Dichotomous traits generally have lower discovery power than continuous traits. The effect size estimates also tend to have larger variance for dichotomous traits. We also found that the effect size estimates from both score and Wald approaches have some small downward bias, consistent with what has been previously reported.

**Extended Data Figure 3.**
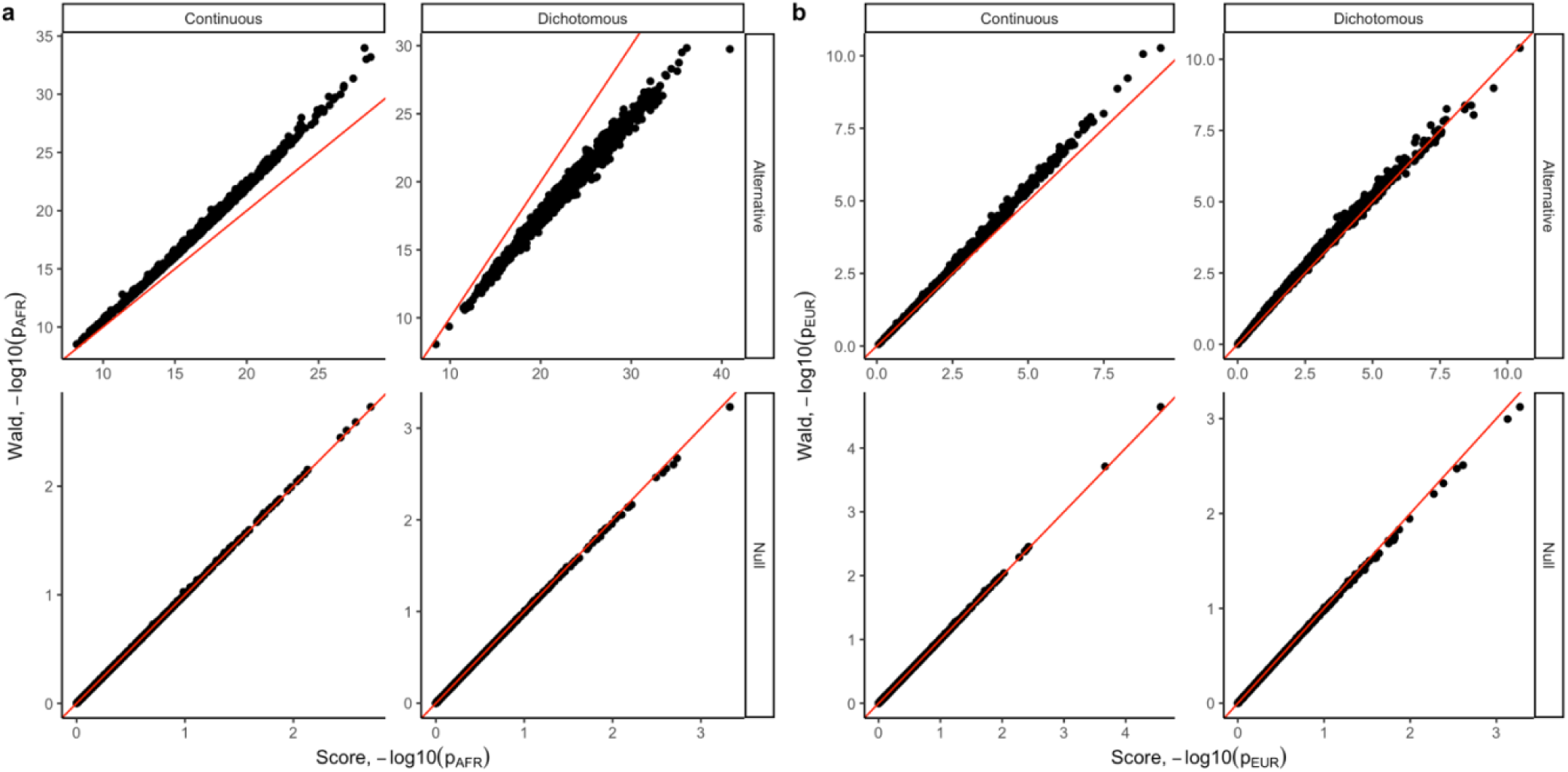
*P* value comparison between the Wald test and the Score test approximation. Under the null hypothesis, *P* values obtained from the Wald test and the Score test were largely consistent. However, the score outcomes are more significant than the Wald for the dichotomous traits and less for continuous traits. (**a**) AFR-specific p values obtained by the Wald test (y-axis) vs score test (x-axis); (**b**) EUR-specific p values obtained by Wald test (y-axis) vs score test (x-axis).

