## Supplementary for "Extending Genome-Wide Association Studies to admixed cohorts with high degrees of relatedness"

Additional information on analysis of the Yale-Penn cohort

*Data processing*: We included 3,063 individuals from the Yale-Penn cohort, which had 2-way admixture between EUR and AFR ancestry. The Yale-Penn cohort includes participants that were recruited for studies of the genetics of drug or alcohol dependence in five eastern U.S. centers, as previously described[1–5]. Subjects were evaluated with the semi-structured assessment for drug dependence and alcoholism (SSADDA). In the present study, we used information regarding alcohol-related traits, such as Total number of drink (TotDrinks), Age of initiation alcohol (AlcAge), and alcohol use disorder (AUD). We defined TotDrinks with the SSADDA question: In your lifetime, what is the largest number of drinks you have ever had in a 24-hour period (including all types of alcohol)? Meanwhile, for AlcAge was defined with How old were you when you took your first real drink of alcohol (not a sip; not at a religious ceremony)? Participants gave written informed consent as approved by the institutional review board at each site. TotDrinks and AlcAge were inverse-rank normalized transformed, AUD was modelled as binary variable.

Yale-Penn was genotyped with the Illumina HumanCore Exome array and imputed using the 1000 Genomes project. We QC-ed the genetic variants such to contain only biallelic SNPs with > 0.5% MAF, > 95% call rate, *P* value for HWE more than $1 \times 10^{-6}$, and INFO scores > 0.8. For construction of the PCs and GRM, we performed LD pruning with PLINK, with the `--indep 50 5 2` flag. King-Robust[6] was used to infer relatedness. We extracted continental AFR and EUR individuals from the Thousand Genomes Project reference data[7] and merged these with the pruned Yale Penn data. Specifying *k*=2, we then ran ADMIXTURE[8] to confirm samples were indeed two-way AFR-EUR admixed. The included samples had a mean proportion of AFR ancestry of 22.22%. KING-Robust analysis reveals that this dataset contains 952 pairs of relatives with a kinship > 0.0625.

We used *Shapeit2* with default parameters for statistical phasing[9], providing continental AFR and EUR from the Thousand Genomes Project as the reference panel. Using the same reference panel, we used *RFMix2* for local ancestry inference[10]. The genetic map files used in Shapeit2 and RFmix2 (b37) were obtained from the SHAPEIT github[11]. To compute PCs and GRM, we performed LD pruning with Plink, and calculated 2 principal components with PC-Air and the GRM with PC-Relate[12–14].

*Tractor-Mix* analysis requires 1) estimated global ancestry or PCs, 2) estimated GRM, 3) partitioned genotype files, and 4) phenotype and covariate files (Extended Data Figure 1). For 1) and 2), we applied PC-Air and PC-Relate on the Yale Penn 2 dataset according to the GENESIS vignette.[15] The top 2 principal components from PC-Air were used for computation with PC-Relate. The estimated GRM from PC-Relate was rescaled (X2 to make the diagonal elements close to 1) and masked such that elements less than 0.05 were set to 0. To obtain partitioned genotype files, we applied the extract_tracts.py step from *Tractor*. The null model was fitted with the `glmmkin()` function from *GMMAT*, providing age, sex, and the top 5 PCs from PC-Air as covariates, and using the sparse PC-Relate as the estimated GRM. To compute the 1 d.o.f. *GMMAT* test, we applied a standard score test (`glmm.score()`) on PLINK format data, with genotypes specified to be non-centered (`center = F`). To compute the 2 d.o.f. *Tractor-Mix* test, we applied `TractorMix.score()`. The output Manhattan plot were visualized with the matplolib python package. When applying *Tractor-Mix* on AUD, we used the default AC = 50 as the threshold for binary traits.

*Results*: In the analysis TotDrinks and AlcAge, we did not find any SNP that reach genome-wide significance due to the modest sample size of this cohort. However, we identify associations for AUD in the GMMAT and *Tractor-Mix* analysis (Supplementary Figure 10). For the GMMAT analysis, we identified 123 SNPs that reach genome-wide significance. For the Tractor-Mix, we identified 2 SNPs in the joint p-value analysis. In the ancestry-specific analysis, Tractor-Mix identified 123 SNPs at genome-wide significance in the AFR ancestry and no genome-wide significance signal in the EUR ancestry, better resolving the origin of association signal.

**Supplementary Figures**

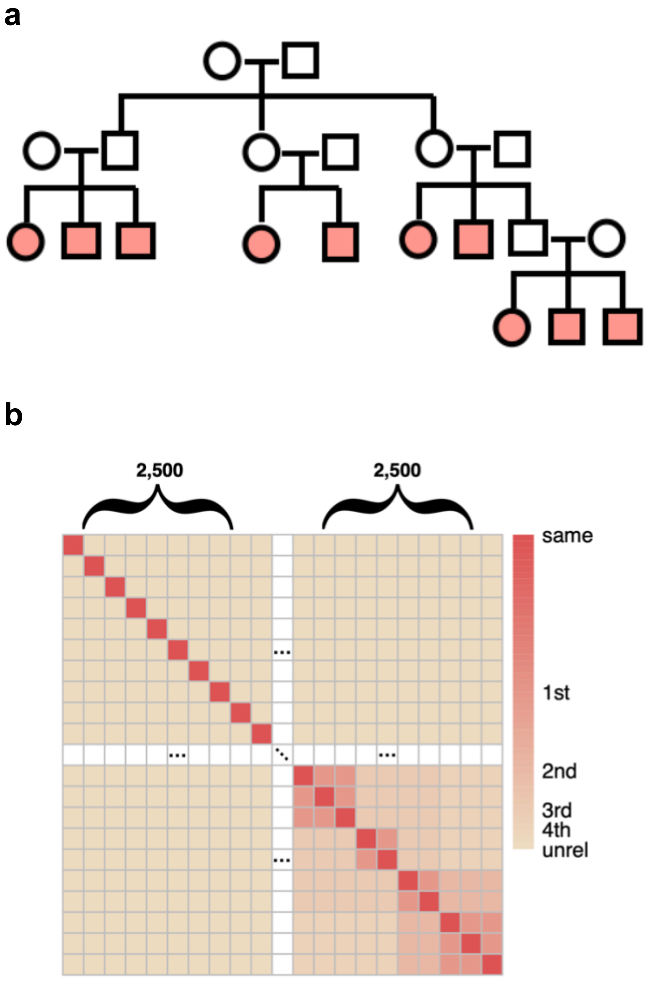


**Supplementary Figure 1**. Pedigree for simulations. We used the pedigree shown in panel **a** to generate relatives for all numerical simulations. The family consists of 20 individuals, with 10 of them (colored in red) included in the association test. In addition to including these related samples in the simulation, we also simulated independent admixed individuals, forming the ultimate kinship matrix shown in panel **b**.

**a**

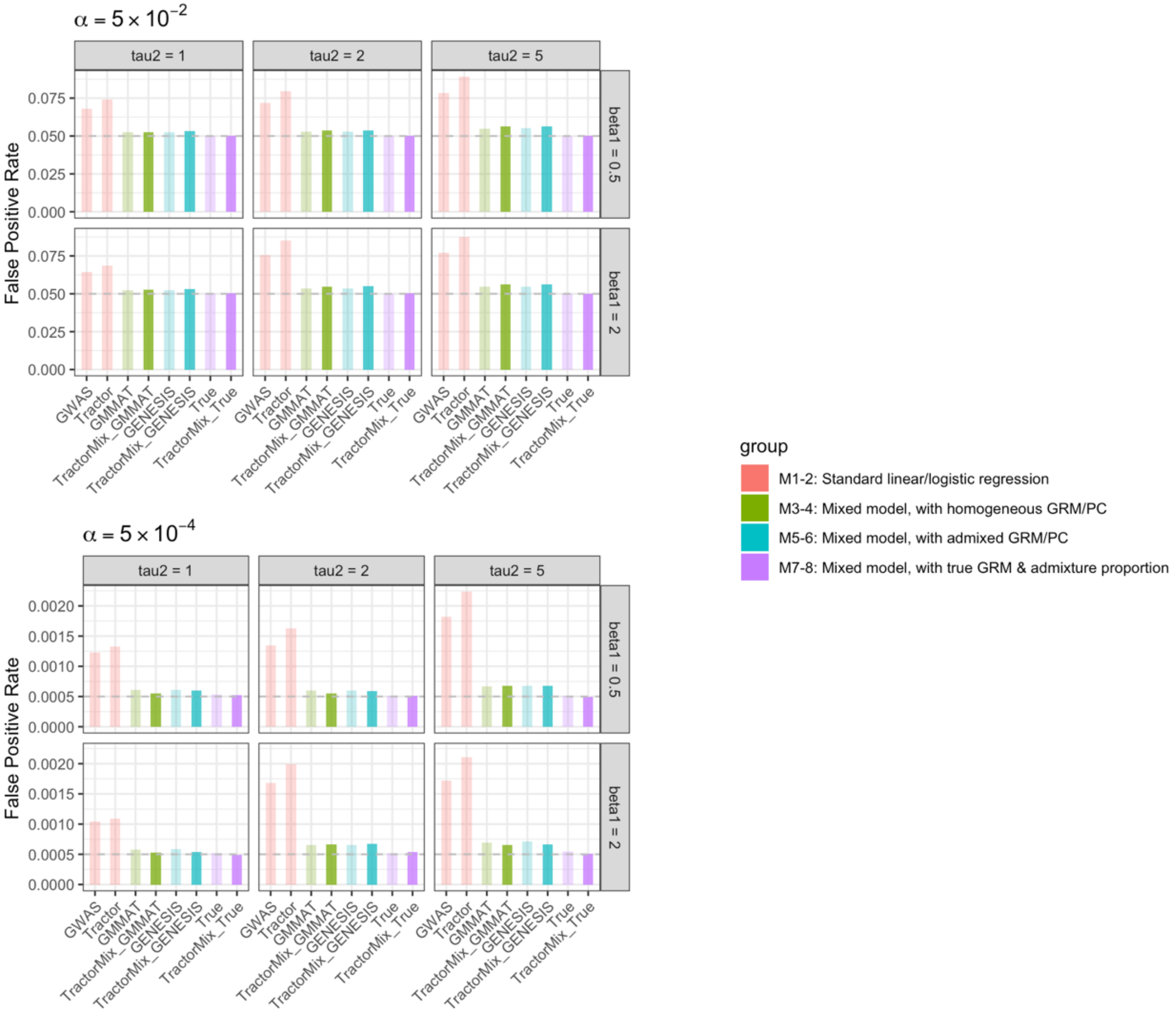

$\alpha = 5 \times 10^{-2}$
tau2 = 1
tau2 = 2
tau2 = 5
beta1 = 0.5
beta1 = 2
False Positive Rate
0.075
0.050
0.025
0.000
GWAS
Tractor
GMMAT
TractorMix_GMMAT
GENESIS
TractorMix_GENESIS
True
TractorMix_True
$\alpha = 5 \times 10^{-4}$
0.0020
0.0015
0.0010
0.0005
0.0000
group
M1-2: Standard linear/logistic regression
M3-4: Mixed model, with homogeneous GRM/PC
M5-6: Mixed model, with admixed GRM/PC
M7-8: Mixed model, with true GRM & admixture proportion

**b**

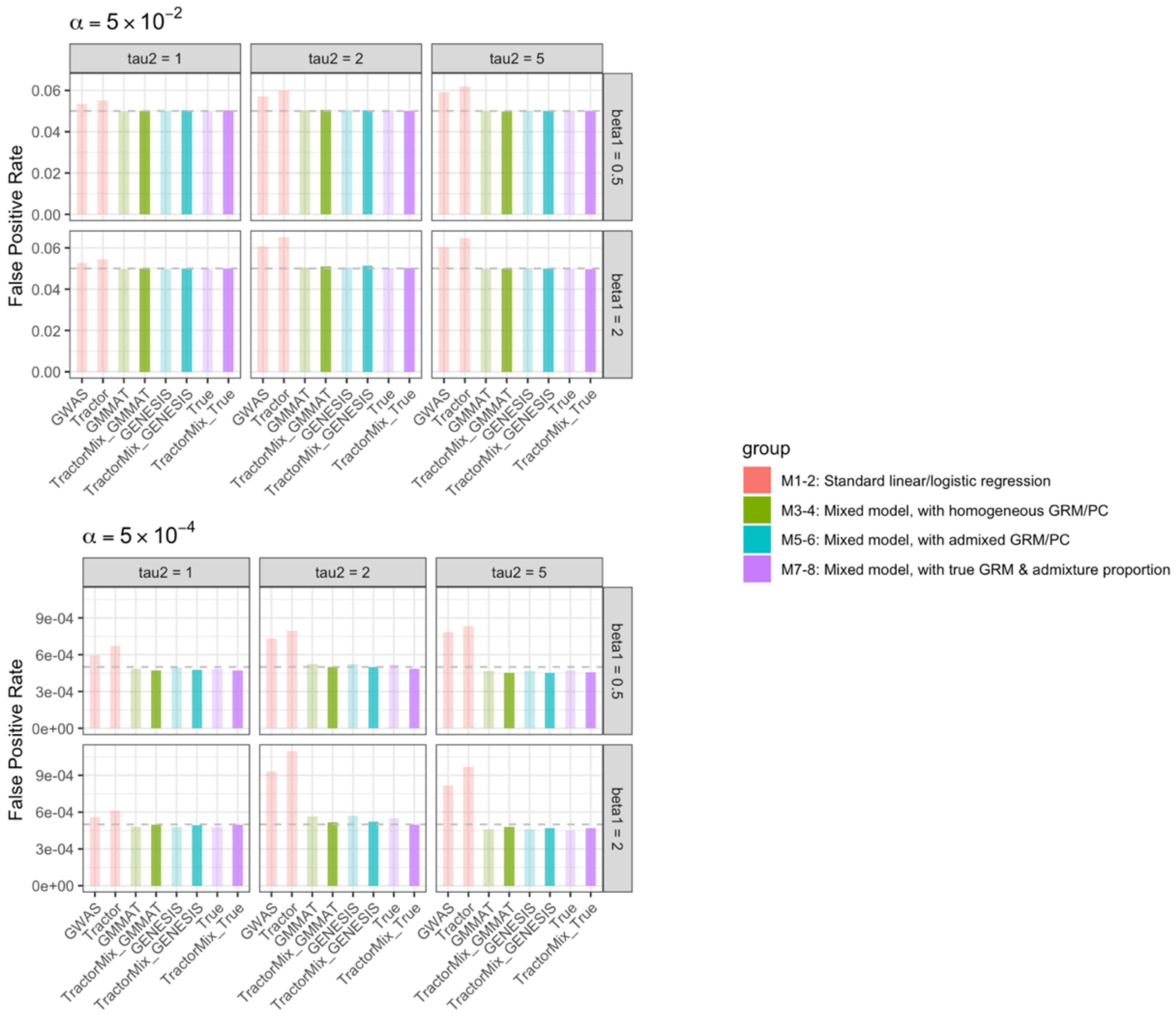


**Supplementary Figure 2**. False positive comparison for (**a**) continuous and (**b**) dichotomous phenotypes. The dichotomous phenotype is modeled with a prevalence of 0.2. We evaluated the false positive rates for $\alpha = 5 \times 10^{-2}$, $5 \times 10^{-4}$ with different parameters $(\beta_1, \tau^2)$ for data generation. $\beta_1$ is the coefficient for admixture proportion, which controls for the degree of population stratification. $\tau^2$ is the coefficient for family relatedness.

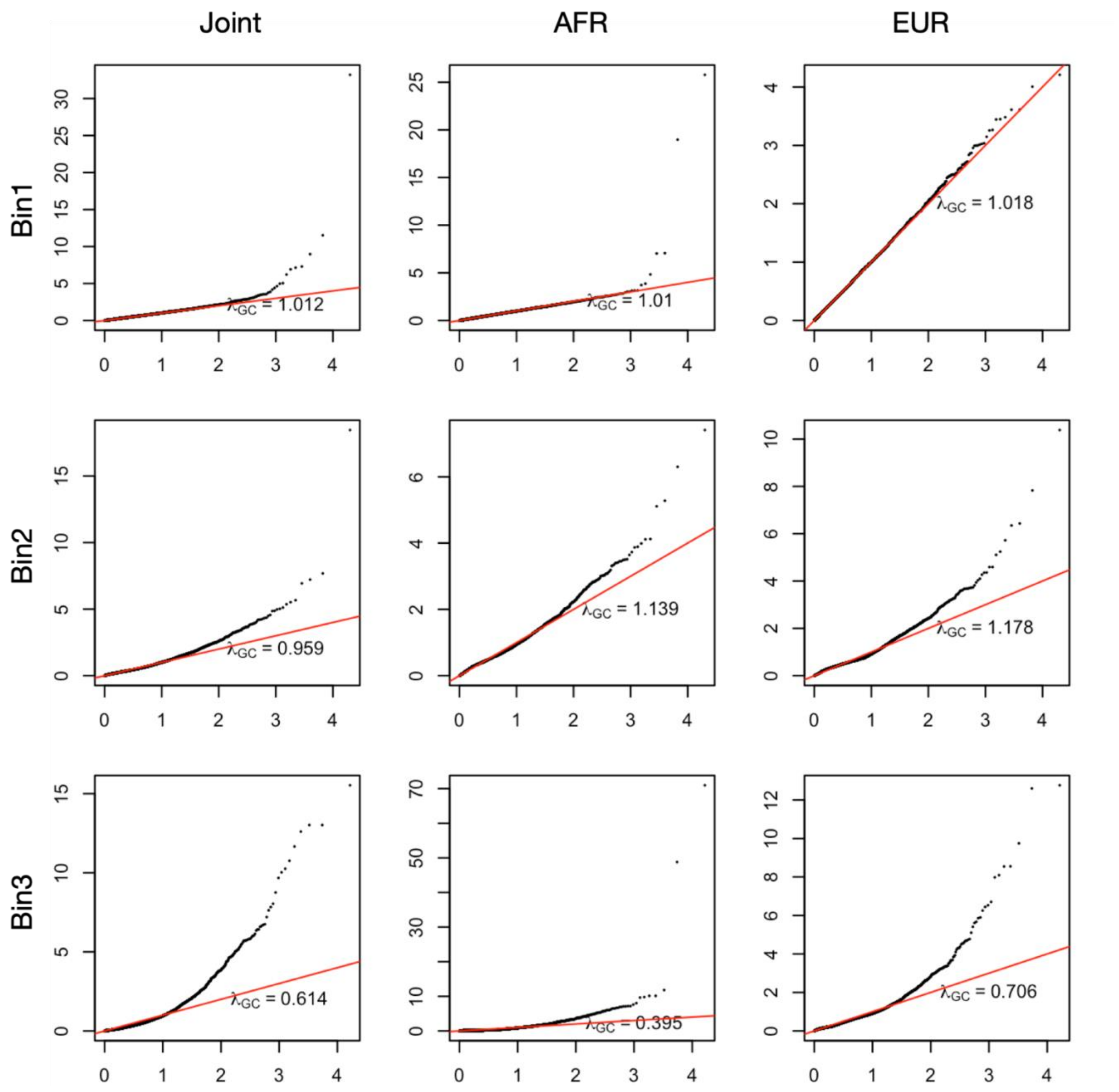


**Supplementary Figure 3.** Increased false positives appear without dropping ancestry-specific dosages with low allele counts. Variants are distributed into 3 bins, with bin1 containing variants with AC > 50 for both ancestries, bin2 containing variants with AC > 10 for both ancestries (excluding bin1), and bin3 containing all remaining variants. *Tractor-Mix* is callibrated for variants in bin1, but not for variants in bin2 or bin3. The QQ plot is generated with UKBB 2-way admixed AFR-EUR samples, using sickle cell anemia as the trait.

**a**

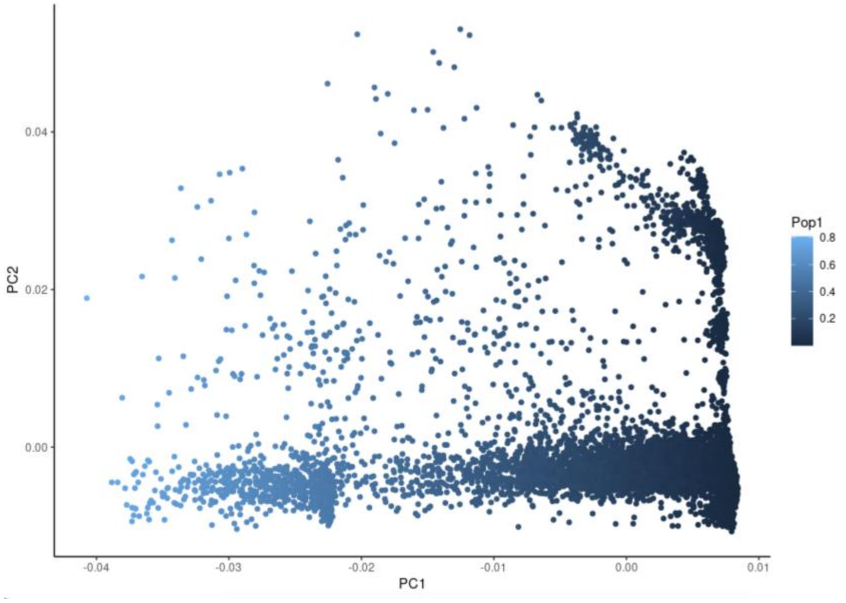


**b**

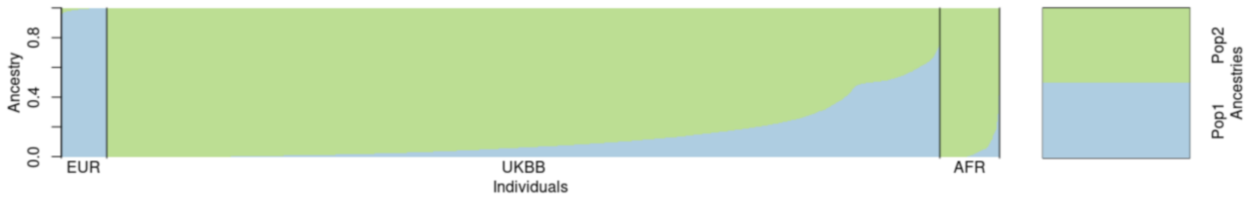


**Supplementary Figure 4.** Global ancestry inference for the utilized UKBB sample. (**a**) Principal components analysis for two-way admixed UKBB AFR-EUR individuals. (**b**) ADMIXTURE plot of the UKBB sample alongside AFR and EUR individuals from the Thousand Genome Project. Note the high degree of admixture and heterogeneity present in the UKBB sample. We found that the admixture porportion obtained from ADMIXTURE is highly consistent with the first principal component, with correlation = 0.998.

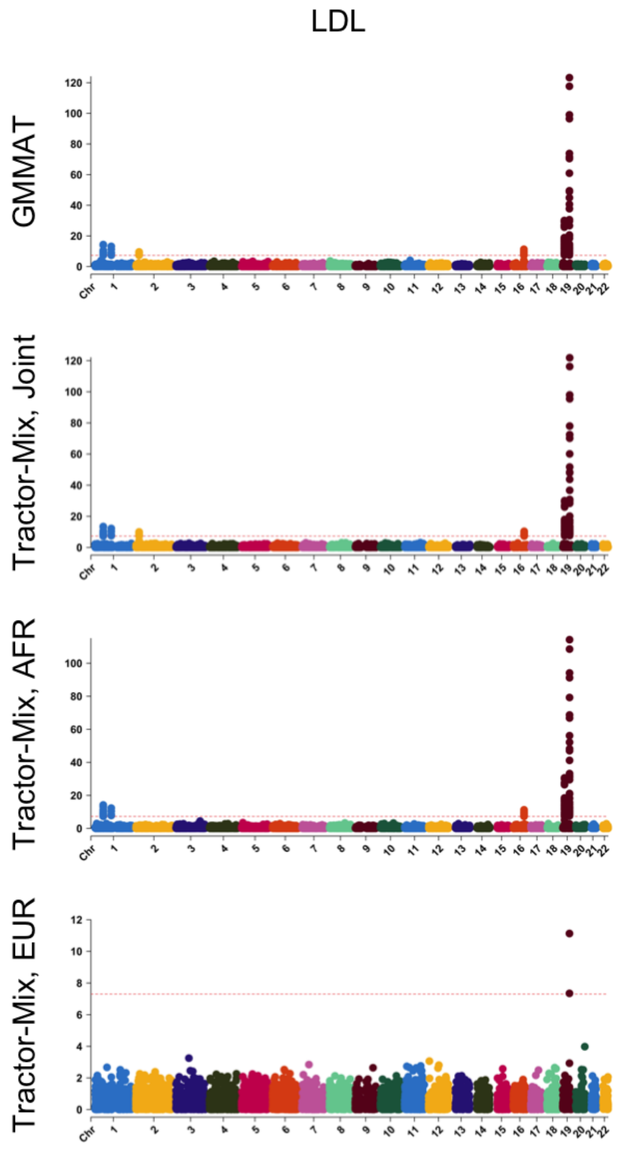


**Supplementary Figure 5.** GWAS results for LDL cholesterol for UKBB two-way admixed AFR/EUR individuals. Rows indicate the statistical model utilized.

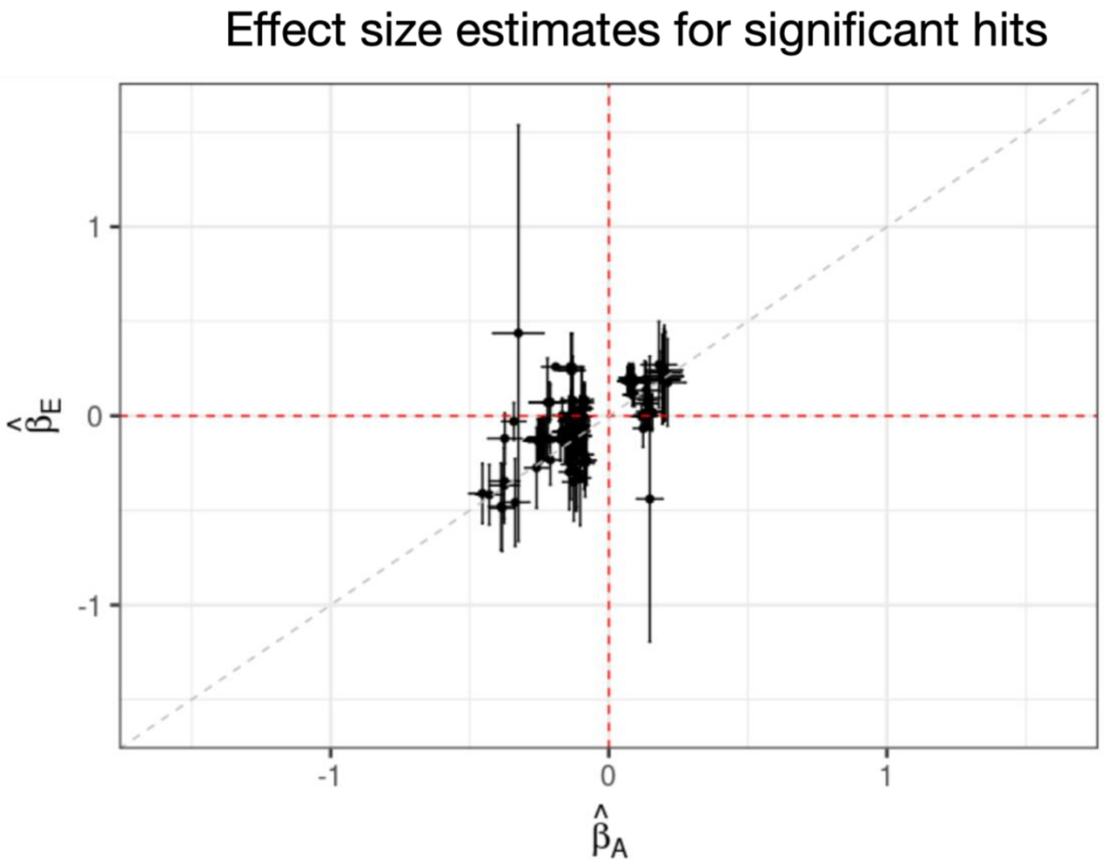


**Supplementary Figure 6.** Ancestry-specific effect size estimates for significant loci of the UK Biobank total cholestral Tractor-Mix GWAS. Only the variants that passed the genome-wide significance threshold were selected for visualization. The x-axis is the effect size estimates of AFR ancestry, while the y-axis is the effect size estimates of EUR ancestry. Point estimates and confidence intervals are shown.

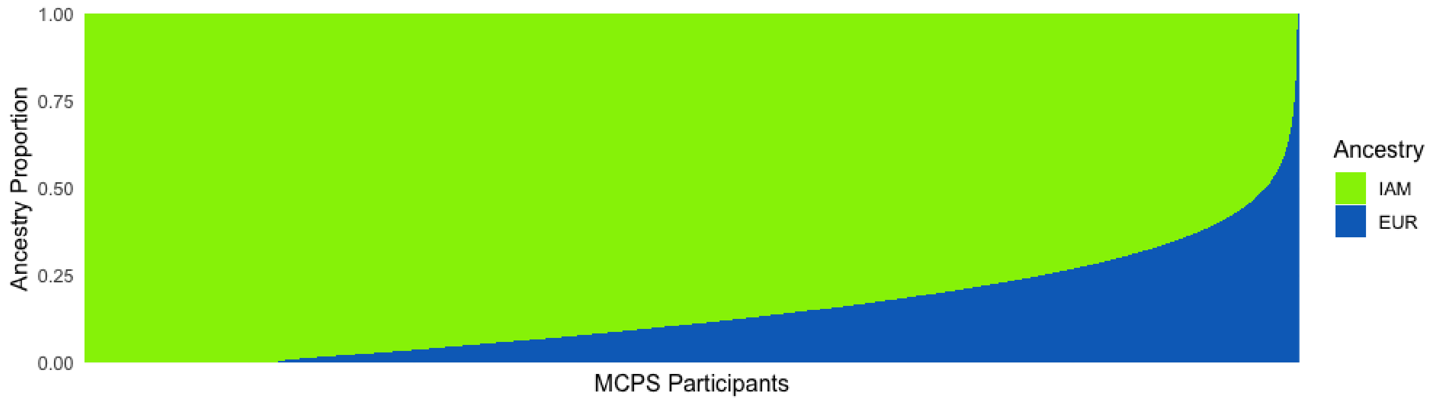


**Supplementary Figure 7**. Ancestry proportions for 33,500 individuals from the MCPS cohort. A four-way model was fit using ADMIXTURE software for a total of 140,829 MCPS participants allowing for genetic inheritance from Indigenous American (IAM), European (EUR), African (AFR), and East Asian (EAS) ancestral populations. Individuals with a combination of AFR and EAS ancestry > 2.5% were excluded from GWAS analyses in order to retain individuals best represented by 2-way admixture from IAM (green) and EUR (blude) ancestries, as shown here.

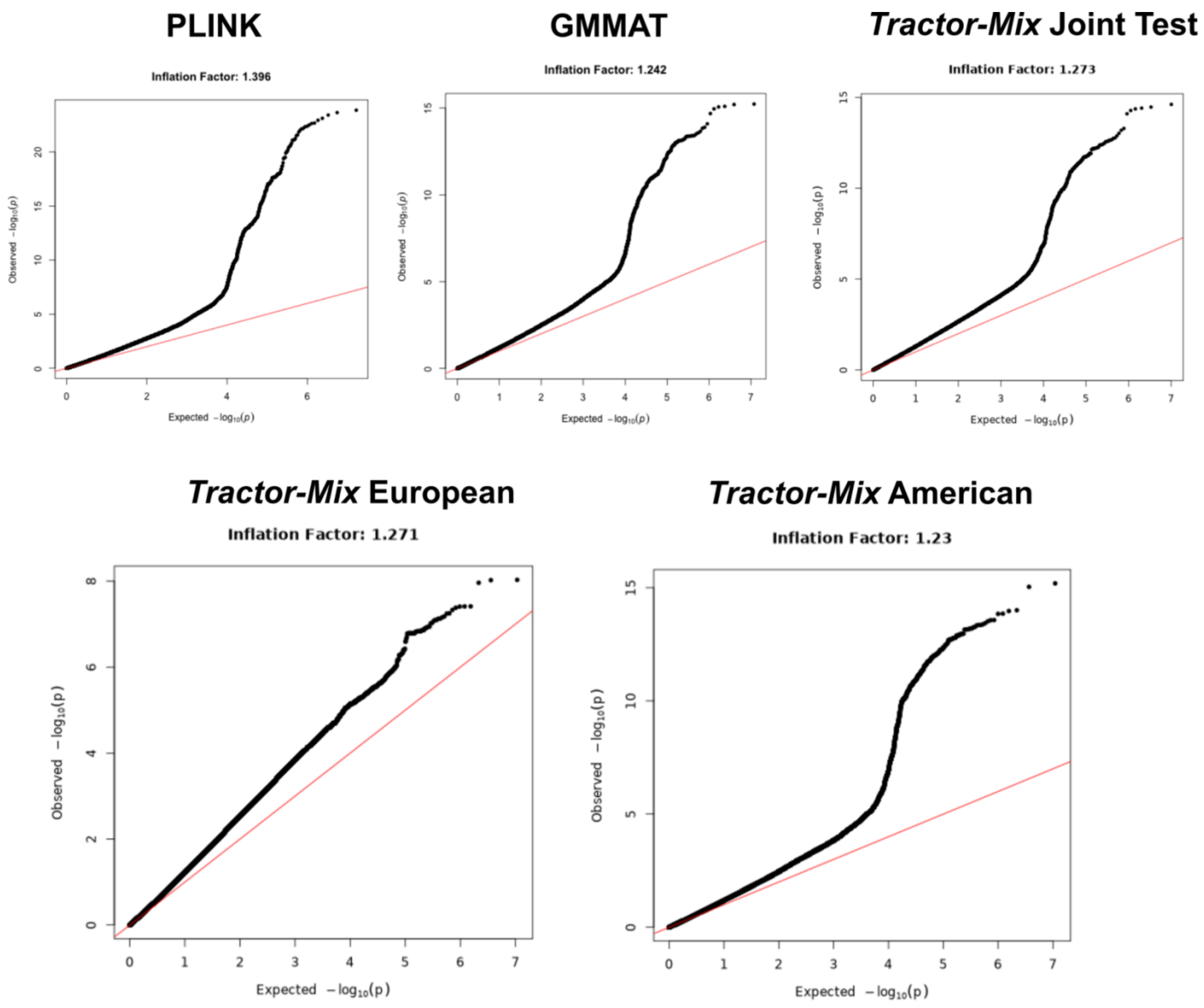


**Supplementary Figure 8.** QQ-plots from GWAS analyses of BMI in MCPS using *PLINK*, *GMMAT*, and *Tractor-Mix*. Genomic control lambda inflation factors are shown for each GWAS.

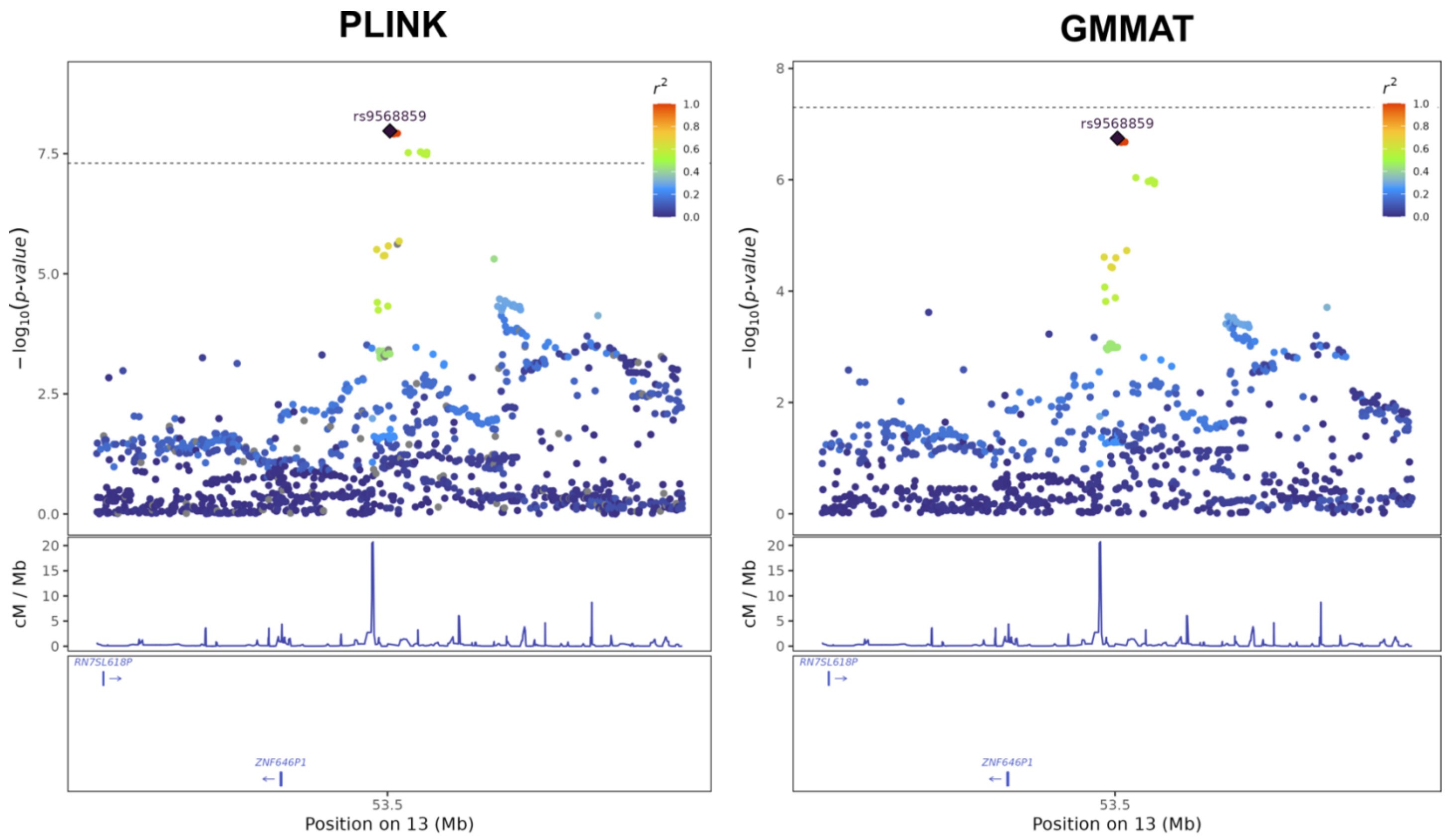


**Supplementary Figure 9**. **Locus plots at the *ZNF646P1* locus from the GWAS of BMI in the MCPS cohort**. Associations for variants mapping within 250kb of the index variant rs9568859 are shown with color corresponding to pairwise LD $r^2$ with respect to this SNP. Results are shown from the analyses using linear regression implemented in *PLINK* or the score test implemented in *GMMAT*.

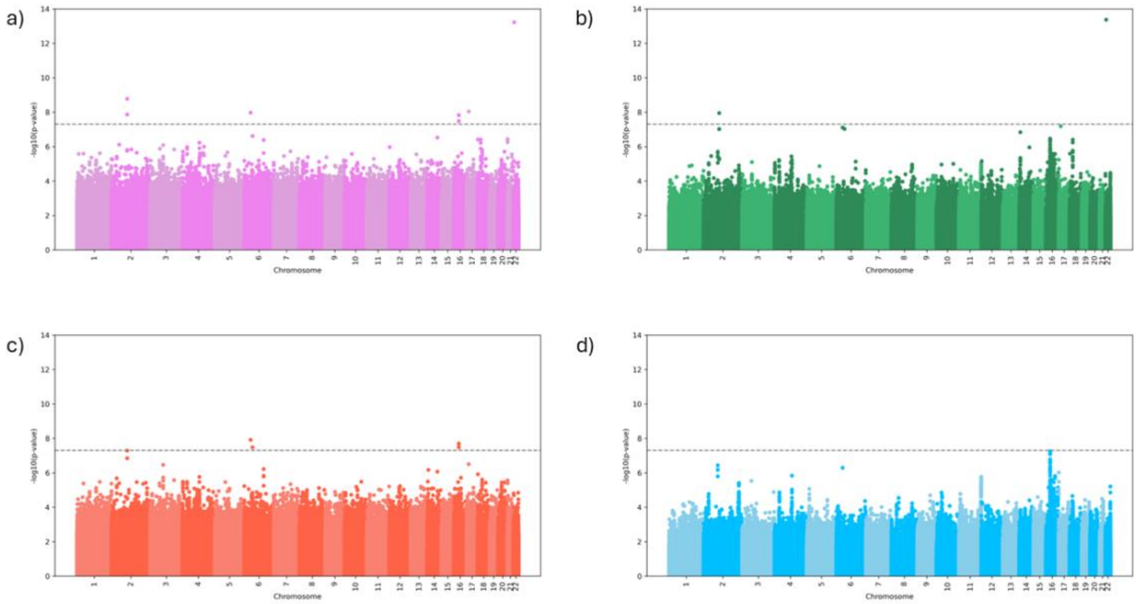


**Supplementary Figure 10.** Manhattan plots for GMMAT and Tractor-Mix for AUD in the Yale Penn cohort. **a**) Manhattan plot for the GMMAT results, **b**) Manhattan plot for the p-values of the joint analysis in Tractor-Mix, **c**) Manhattan plot for the AFR specific p-values for the Tractor-Mix, and **d**) Manhattan plot for the EUR specific p-values for the Tractor-Mix.

### Supplementary Tables

| | Threshold | Joint *P* value | AFR *P* value | EUR *P* value |
| --- | --- | --- | --- | --- |
| **Continuous (120M)** | 5e-2 | 5.0e-2 | 5.0e-2 | 5.0e-2 |
| | 5e-4 | 4.9e-4 | 5.0e-4 | 4.9e-4 |
| | 5e-6 | 4.7e-6 | 4.6e-6 | 4.8e-6 |
| **Dichotomous (22M)** | 5e-2 | 5.0e-2 | 5.0e-2 | 5.0e-2 |
| | 5e-4 | 4.9e-4 | 4.9e-4 | 5.0e-4 |
| | 5e-6 | 4.8e-6 | 5.2e-6 | 4.3e-6 |

**Supplementary Table 1**. False positive evaluation for continuous and dichotomous traits. We evaluated the false positive rate for both continous and dichotomous phenotypes, providing the true data generating covariate and kinship matrix. For each iteration, we compute the joint *P* value (2 d.o.f.), as well as approximating ancestry-specific *P* values (AFR and EUR). We assess false positives using different thresholds of $\alpha = 5 \times 10^{-2},\ 5 \times 10^{-4}, \text{and } 5 \times 10^{-6}$, which required increasing number of iterations for computation. Given computational considerations, we computed ~120M *P* values for the continuous phenotype and ~22M *P* values for the dichotomous phenotype.

**a**

| CHR:POS | SNP ID | MAF (AFR) | MAF (EUR) | BETA (AFR) | PVAL (AFR) | BETA (EUR) | PVAL (EUR) |
|---|---|---|---|---|---|---|---|
| 19:45412079 | rs7412 | 0.120 | 0.068 | -0.45 | 1.3E-78 | -0.41 | 2.6E-07 |
| 1:55518622 | rs45613943 | 0.303 | 0.051 | -0.13 | 1.4E-15 | -0.25 | 6.6E-03 |
| 16:72217018 | rs61483465 | 0.186 | 0.135 | 0.14 | 1.2E-12 | 0.09 | 9.8E-02 |
| 2:21295227 | rs62122515 | 0.189 | 0.338 | 0.08 | 2.3E-05 | 0.20 | 1.6E-07 |

**b**

| CHR:POS | SNP ID | MAF (AFR) | MAF (EUR) | BETA (AFR) | PVAL (AFR) | BETA (EUR) | PVAL (EUR) |
|---|---|---|---|---|---|---|---|
| 19:45412079 | rs7412 | 0.120 | 0.068 | -0.45 | 1.3E-78 | -0.41 | 2.6E-07 |
| 1:55518622 | rs45613943 | 0.303 | 0.051 | -0.13 | 1.4E-15 | -0.25 | 6.6E-03 |
| 16:72217018 | rs61483465 | 0.186 | 0.135 | 0.14 | 1.2E-12 | 0.09 | 9.8E-02 |
| 2:21295227 | rs62122515 | 0.189 | 0.338 | 0.08 | 2.3E-05 | 0.20 | 1.6E-07 |

**Supplementary Table 2.** Comparing *P* values for the leading SNPs of genome-wide significant locus for total cholestral. (**a**) Effect size estimates and *P* values for the Wald test. (**b**) Effect size estimates and *P* values for the score test. The effect sizes and p-values are identical after round to 2 significant figures.
